# Exploring test retest reliability and longitudinal stability of digital biomarkers for Parkinson’s disease in the m-Power dataset

**DOI:** 10.1101/2020.12.16.20247122

**Authors:** Mehran Sahandi Far, Simon B. Eickhoff, María Goñi, Juergen Dukart

**Author notes:** Contributed equally (María Goñi, Juergen Dukart). **To whom correspondence should be addressed: Juergen Dukart, PhD**, Institute of Neuroscience and Medicine, Brain & Behaviour (INM-7), Research Centre Jülich, Jülich, Germany.

## Abstract

**Background:** Digital biomarkers (DB) as captured using sensors embedded in modern smart devices are a promising technology for home-based symptom monitoring in Parkinson’s disease (PD).

**Objective:** Despite extensive application in recent studies test-retest reliability and longitudinal stability of DB has not been well addressed in this context. We utilized the large-scale m-Power dataset to establish the test-retest reliability and longitudinal stability of gait, balance, voice and tapping tasks in an unsupervised and self-administered daily life setting in PD patients and healthy volunteers.

**Methods:** Intraclass Correlation Coefficients (ICC) were computed to estimate the test-retest reliability of features that also differentiate between PD and healthy volunteers. In addition, we tested for longitudinal stability of DB measures in PD and HC as well as for their sensitivity to PD medication effects.

**Results:** Among the features differing between PD and HC, only few tapping and voice features had good to excellent test-retest reliabilities and medium to large effect sizes. All other features performed poorly in this respect. Only few features were sensitive to medication effects. The longitudinal analyses revealed significant alterations over time across a variety of features and in particular for the tapping task.

**Conclusions:** These results indicate the need for further development of more standardized, sensitive and reliable DB for application in self-administered remote studies in PD patients. Motivational, learning and other confounds may cause a variation in performance that needs to be considered in DB longitudinal applications.

## Introduction

Parkinson’s disease (PD) is primarily characterized by motor symptoms including tremor at rest, rigidity, akinesia, and postural instability [1]. Although standard in-clinic assessments such as the Unified Parkinson’s Disease Rating Scale (UPDRS) are popular, they are influenced by inter-rater variability by relying on self-reporting by patients and care givers or clinician judgement [2]. In addition, they are costly and limited with respect to observation frequency.

The emergence of new technologies has led to a variety of sensors (i.e. acceleration, gyroscope, GPS, etc.) embedded in smart devices of daily use (i.e., smartphone, smartwatch). Such sensor data alongside with other digital information recorded passively or when executing prespecified tasks may provide valuable insight into health-related information. Such applications are now commonly referred to as digital biomarkers (DB) [3]. DB being collected frequently over a long period of time can provide an objective, ecologically valid and more detailed understanding of the inter- and intra-individual variability in disease manifestation in daily life.

Numerous DB have been proposed for PD diagnosis as well as to establish a link with clinical rating scales such as UPDRS and to quantify disease severity or intervention effect [4–8]. Despite these various proof of concept studies, many technical challenges with respect to DB deployment remain unaddressed. DB measures are prone to large variation caused by technical and procedural differences including but not limited to placement/orientation, recording frequency of the devices and environmental and individual variation, i.e. due to motivation, medication or other aspects [9– 11]. Other factors such as the effect of users’ familiarity with technology and the impact of learning on the performance of measured DB in remote PD assessment are other important sources of variation that have not been addressed so far. All of these factors may limit the sensitivity and reliability of DB measurements for any of the above PD clinical applications. DB longitudinal variation is therefore an important attribute that should be quantified and addressed. The reliability of DB assessment has been broadly studied for gait, balance, voice and tapping data [12–17]. However, the existing studies typically focused on a single or few aspects of PD and most of them established the test-retest reliability in a standardized clinical setting limiting the translatability of their findings to at-home applications. Among studies that evaluated DB assessments for remote monitoring of PD, only one reported the test-retest reliability [5]. No PD studies systematically evaluated the test-retest reliability and longitudinal sensitivity of DB in a fully unsupervised and self-administered PD longitudinal setting.

Here aimed to address these open questions on the performance of DB measures in PD when collected in a self-administered setting in daily life. For this, we first performed a comprehensive literature search identifying 773 DB features reported in previous studies to cover PD-related alterations in gait characteristics, tremor, postural instability, voice and finger dexterity. We evaluated the longitudinal stability and test re-test reliability of these features as collected using four commonly applied PD tasks (gait, balance, voice and tapping) in daily life in a large cohort of self-reported PD patients and healthy controls (HC), the m-Power study [18]. In addition, we evaluated their sensitivity to learning and medication effects.

## Methods

### Study cohort

We utilized smartphone-based data from a large-scale longitudinal observational cohort collected in the first six month of m-Power study [18]. The enrolment was open to adult participants who own an iPhone, are living in the US and are comfortable with English to read the instructions in the app. Participants were asked to download the application and complete a one-time demographic survey during registration. Demographic data includes but not limited to age, sex, health history, and previous PD clinical diagnosis. They also were asked to fill out a survey with selected questions from the movement disorder society’s UPDRS, as well as, PDQ-8. All the participants were suggested to complete each task (walking, tapping, voice, and memory) up to three times a day for up to six months. In addition, self-reported PD patients were asked to complete the task before medication, after medication and at another time when they are feeling at their best.

Ethical oversight of the m-Power study was obtained from Western Institutional Review Board. Prior to signing an electronically rendered traditional informed consent form, prospective participants had to pass a five-question quiz evaluating their understanding of the study aims, participant rights, and data sharing options. After completing the e-consent process and electronically signing the informed consent form, participants were asked for an email address to which their signed consent form was sent and allowing for verification of their enrolment in the study. Participants were given the option to share their data only with the m-Power study team and partners (‘share narrowly’) or to share their data more broadly with qualified researchers worldwide, and had to make an active choice to complete the consent process (no default choice was presented). The data used in our study consist of all individuals who chose to have their data shared broadly.

### Data Pre-processing

The m-Power dataset is assessed outside of clinical environment with severely limited quality control and supervision. All information including the health history, diseases diagnosis, duration, treatment and survey outcomes are self-reported. To address these, we excluded participants, who did not specify their age, sex, or information on professional diagnosis (if they belong to the PD or HC group). The participants are assigned to PD or HC group according to their answer to the question “Have you been diagnosed by a medical professional with Parkinson disease?”. There was a significant difference in the age and sex distribution between HC and PD subjects, particularly age slanted toward younger and male individuals in HC. To reduce the impact of age, we restricted the age range for our analysis to between 35 and 75 years. The resulting number of available assessments per subject is displayed in Table 1.

**Table 1.**
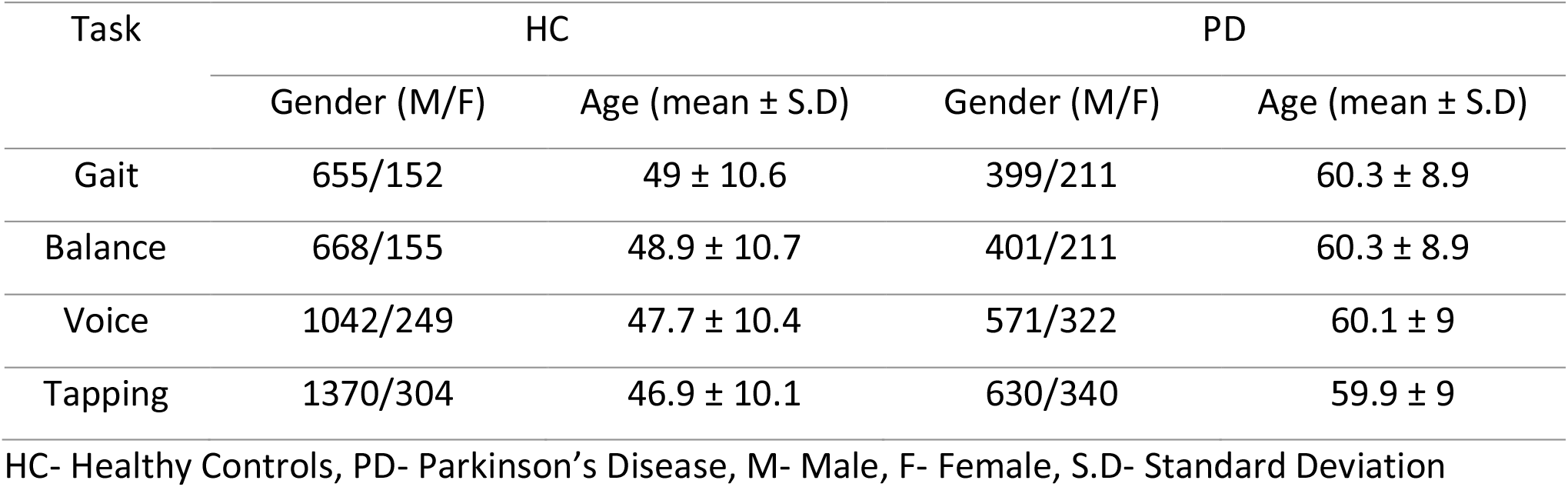
Characteristics of study cohorts after data cleaning.

### Feature extraction

To identify features that are commonly used for the walking, voice and tapping tasks for PD applications, we performed a comprehensive literature search in PubMed with the following terms ((Parkinson’s disease) AND (walking OR gait OR balance OR voice OR tapping) AND (wearables OR smartphones)). Based on this search, we identified an overall of 773 features related to gait (N=423), balance (N=183), finger dexterity (N=43), and speech impairment (N=124). All of these features were computed for the m-Power study. A detailed explanation of the extracted features including the respective references is provided in Table S1-S4. For features sharing the same variance (high pair-wise correlation: Spearman rho>.95), only one of the features was selected randomly for further analyses to reduce the amount of redundant information for each task.

#### Gait and balance

Impairments in gait speed, stride length, and stride-time-variability are common changes that are linked to PD [19–22]. Instability in postural balance is also considered as one of the well-reported characteristics associated with PD [14,23–25]. Both were assessed by a walking task. The gait part consisted of 20 steps walking in a straight line, followed by the balance part of a 30 seconds stay still period. Given a heterogeneity of gait signal lengths across subjects, we used a fixed length signal of 10 seconds and selected data from participants who met this criterion which resulted in 28150 records from 1417 unique participants. In addition to the accelerometer signals (x, y and z), their average, the step series, position along the three axes by double integration, and velocity and acceleration along the path were used for feature extraction [26,27] (Table S1). For balance, we used a 15 seconds time-window trimming the first five and the last 10 seconds of the 30 second records to reduce the noise due to the between-task transition period, resulting in 29050 records from 1435 unique participants. Feature extraction covered signals related to tremor acceleration predicted to fall in the 4-7 Hz band and postural acceleration (non-tremor) falling to the 0-3.5 Hz band [28] (Table S2).

#### Voice

PD may also affect breathing and results in alterations in speech and voice. Reduced volume, hoarse quality, and vocal tremor are commonly reported for PD using voice analysis [15,29,30]. In this task, participants were saying “aaaah” for about ten seconds. For voice, 49676 records were selected belonging to 2184 unique participants. Voice features were computed from fundamental frequency, amplitude and period signals trimming the first and the last two seconds of the ten second interval

#### Tapping

Impairment in finger dexterity is another symptom associated with PD [31,32]. In the m-Power study, participants were asked to tap as fast as possible for 20 seconds with the index and middle fingers on the screen of their phone (positioned on a flat surface). Screen pixel coordinate (x, y) and timestamp of taped points plus acceleration sensor data were collected for this task. Overall, 55894 recordings were selected belonging to 2644 unique participants. Features were computed based on the inter-tapping distance and interval (Table S4).

### Statistical analysis

For features to be considered usable for biomarker purposes in longitudinal studies several criteria are important including among other sensitivity to disease symptoms, good test-retest reliability and robustness against the effects of learning and other longitudinal confounds. To address these criteria, we adopted a step-wise statistical procedure.

As DB measures are frequently not normally distributed, Mann-Whitney U test were used to identify all features that significantly differ between PD and HC at the first administration (baseline) (p<0.05). Effect sizes Cohen’s d were computed for these features to provide an estimate of the magnitude of differentiation between PD and HC.

Next, Intraclass Correlation Coefficients (ICC, type 1-1) were used to determine the test-retest reliability of features showing a significant differentiation between PD and HC. ICC Values of 0.0–0.40 were considered as poor, 0.40–0.59 as fair, 0.60–0.74 as good, and 0.75–1.00 to be excellent [33]. To assess the reliability of each feature, ICCs were computed for different time points vs baseline (one hour (0-6 hours), one day (calendric day), one week (7 calendric days) or one month apart (30 calendric days)), as well as for different repeats versus baseline (baseline versus 2^nd^, 3^rd^, 4^th,^ and 5^th^ repeat). We then selected the top 10 features with the highest median ICCs for each group (PD, HC) and tested for their longitudinal stability over time. For this, we computed repeated measure analyses of variance (rm-ANOVA) using a mixed factorial design with a between-subject factor diagnosis and a within-subject factor repetition (1^st^, 2^nd^, 3^rd^, 4^th,^ and 5^th^) including their interaction. Patients who had at least four repetitions after baseline (463 for gait, 597 for balance, 1085 for voice and 1333 for tapping) were included in these analyses. Lastly, we assessed the impact of PD medication by computing rm-ANOVAs in the PD group with the within-subject factor medication (i.e. before, after, and at best). Patients who had at least one marked task for each of the three PD medication conditions (i.e. before, after, and at best) were included in treatment effect analysis (188 for gait, 189 for balance, 280 for voice and 338 for tapping).

## Results

### Differentiation between PD and HC

First, we aimed to restrict the test-retest reliability analyses of the initial 773 features to those which significantly differ between PD (N=610 to 970 depending on the task, Table 1) and HC (N=807 to 1674). For this, we performed group comparisons for all computed features for gait, balance, voice and tapping tasks. For gait 66 out of 423, for balance 59 out of 183, for voice 60 out of 124 and for tapping 25 out of 43 features differed significantly (p<0.05) between PD and HC at baseline with small (gait and balance) to medium effect sizes for gait, balance and voice and small to large effect sizes for the tapping task (Figure 1 and Table S6-S9).

**Figure 1.**
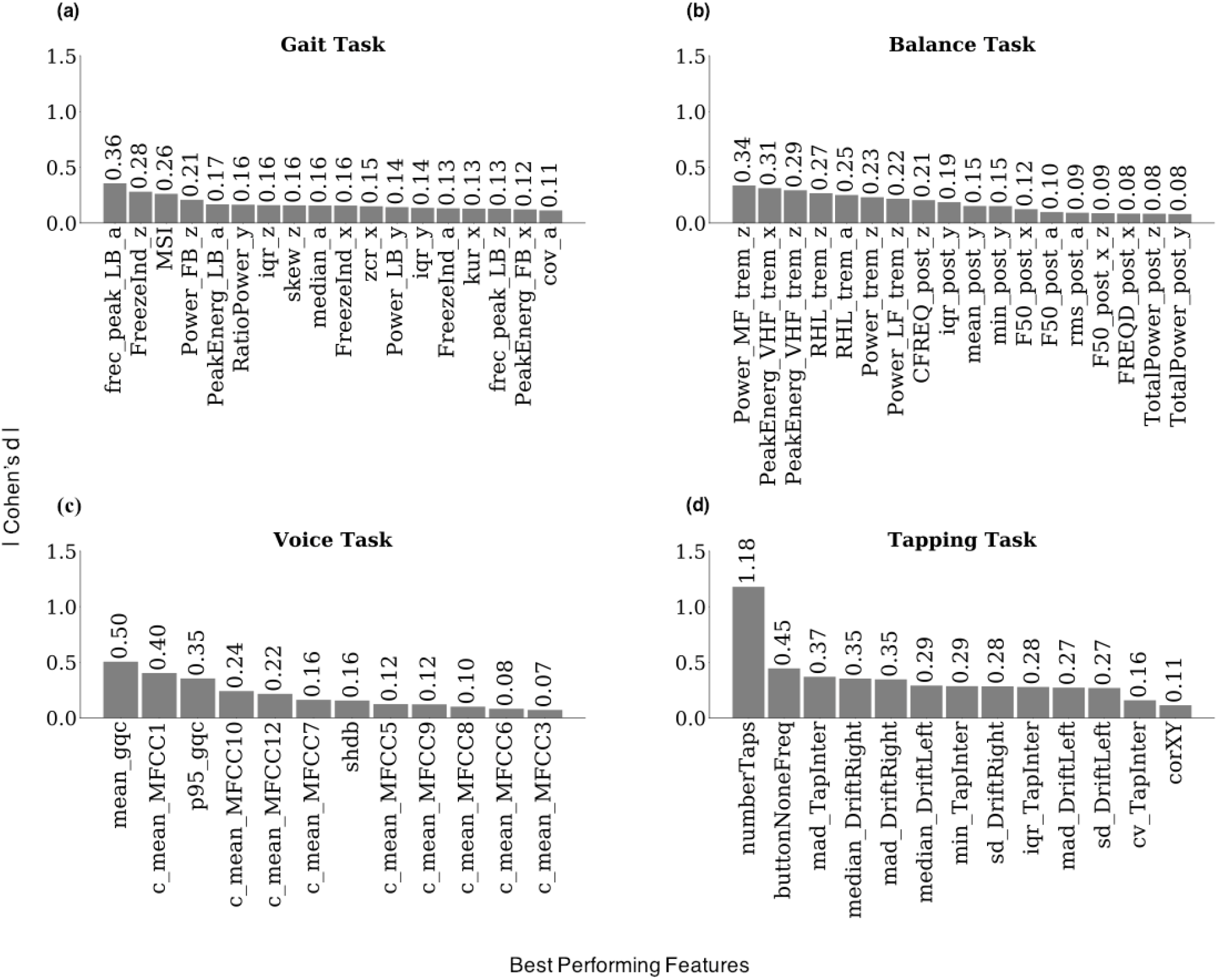
Effect size (Cohen’s d). Best-performing features selected from different time points and repetitions. **(a)** Gait Task: FreezeInd- Freeze Index, PeakEnerg- Peak of Energy, skew- Skewness, MSI- Mean Stride Interval, RatioPower - Sum of the Power in the Freezing and Locomotor Band, frec_peak- Frequency at the Peak of Energy, iqr- Interquartile Range, cov- Coefficient of Variation, zcr- Zero-Crossing Rate, kur-Kurtosis, LB- Locomotor Band, FB- Freezing Band, a- Accelerometer average signal, x- Accelerometer Mediolateral Signal, y- Accelerometer Vertical Signal, z- Accelerometer Anteroposterior Signal, **(b)** Balance Task: PeakEnergy - Peak of energy, TotalPower- Energy between 15-3.5 Hz, Power- Energy between 3.5-15Hz, rms- Root Mean Square, F50- Frequency Containing 50% of Total Power, FRQD- Frequency of Dispersion of the Power Spectrum, iqr- Interquartile Range, min- Minimum Value, CFREQ- Centroidal Frequency, RHL- Ratio Between Power in High Frequency and Low Frequency, MF- Medium Frequency (4-7Hz), VHF- Very High Frequency (>7Hz), HF- Hight Frequency (>4Hz), LF- Low Frequency (0.15-3.5Hz), trem- Tremor, post- Postural, a- Accelerometer Average Signal, x- Accelerometer Mediolateral Signal, y- Accelerometer Vertical Signal, z- Accelerometer Anteroposterior Signal, Hz- Hertz, **(c)** Voice Task: c_mean- Mean of the MFCCs Coefficients, log-Energy of the Signal and the First and Second Derivatives of the MFCCs, MFCC- Mel Frequency Cepstral Coefficients, gqc- Glottis Quotient Close, p95- 95th Percentile, shbd- Shimmer, **(d)** Tapping Task: iqr- Interquartile Range, TapInter- Tap Interval, buttonNoneFreq: Frequency of Tapping Outside the Button, numberTaps- Number of Taps, DriftRight- Right Drift, corXY- Correlation of X and Y Positions, DriftLeft- Left Drift, mad- Median Absolute Deviation, min- Minimum, cv- Coefficient, Sd- Standard Deviation.

### Test-retest reliability

Next, we identified separately for PD and HC the top 10 features with highest median test retest reliability (as measured using intraclass correlation coefficients – ICC) across different time points (one hour, one day, one week or one month apart) and repetitions (all subjects with 5 repetitions of the task) (Table S6-S9). This procedure resulted in 12 to 15 features (including shared ones) being selected for each task (Figure 2, S1 and S2). ICC analyses revealed poor to good test-retest reliability for these most reliable features from the gait and balance tasks and good to excellent reliability for features from voice and tapping tasks (Figure 2). The average ICC across the best performing features selected from different repetitions was lower at the fifth repetition compared to the first, it dropped from .11 to .09 for the gait task, from .21 to .13 for the balance task, from .39 to .24 for the voice task and from .3 to .23 for the tapping task. The average ICC across the best performing features selected from different time points was also lower at one-month compared to one-hour, it dropped from .13 to .07 for the gait, from .2 to .12 for the balance, from .33 to .26 for the voice and from .32 to .19 for the tapping task.

**Figure 2.**
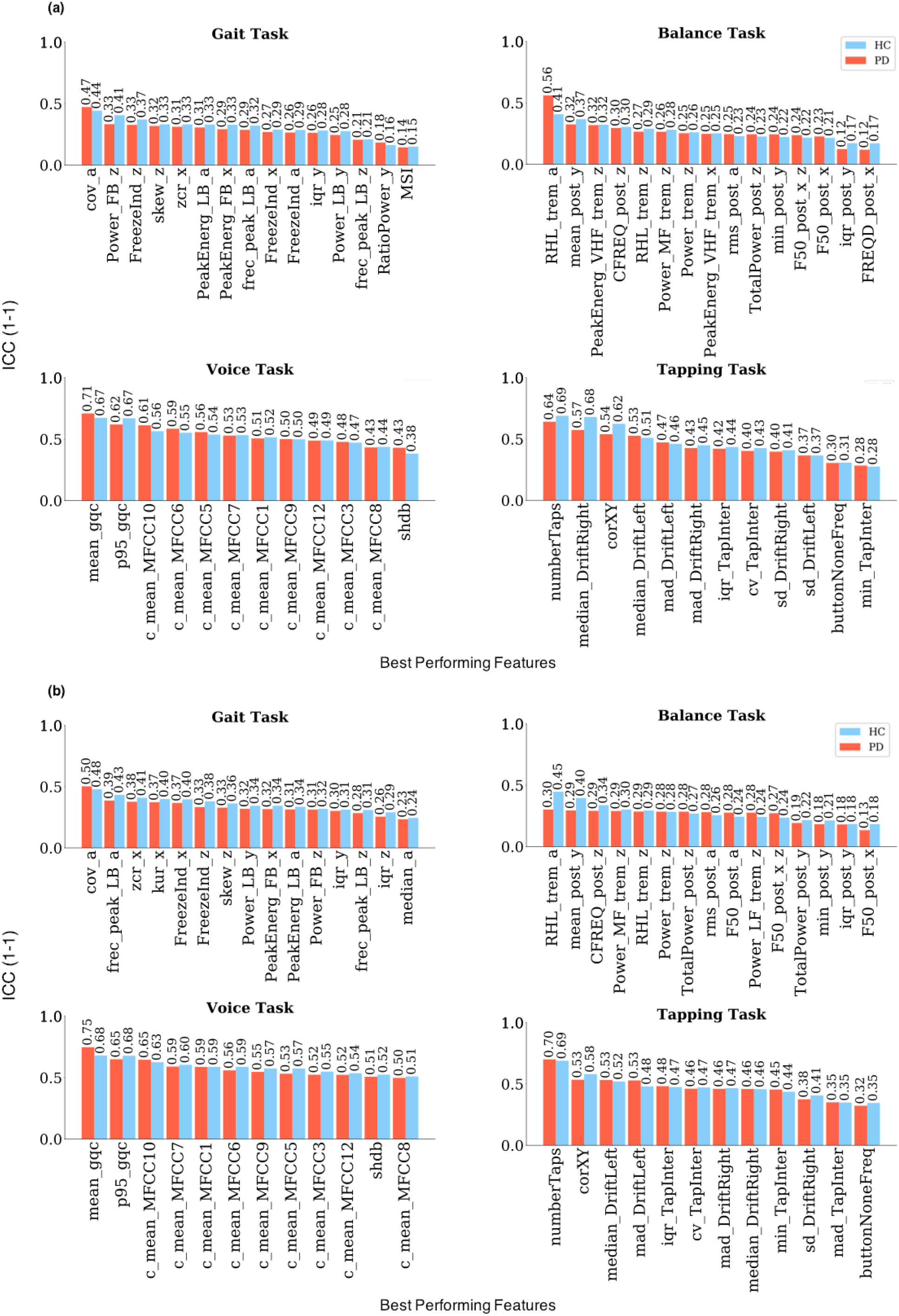
Median ICCs for the best performing features. **(a)** Median ICCs across different time points for the best performing features. **(b)** Median ICCs across different repetitions for the best performing features. Gait Task: FreezeInd- Freeze Index, PeakEnerg- Peak of Energy, skew- Skewness, MSI- Mean Stride Interval, RatioPower - Sum of the Power in the Freezing and Locomotor Band, frec_peak- Frequency at the Peak of Energy, iqr- Interquartile Range, cov- Coefficient of Variation, zcr- Zero-Crossing Rate, kur- Kurtosis, LB- Locomotor Band, FB- Freezing Band, a- Accelerometer Average Signal, x- Accelerometer Mediolateral Signal, y- Accelerometer Vertical Signal, z- Accelerometer Anteroposterior Signal, Balance Task: PeakEnergy - Peak of Energy, TotalPower- Energy between 15-3.5 Hz, Power- Energy between 3.5-15Hz, rms- Root Mean Square, F50- Frequency Containing 50% of Total Power, FRQD- Frequency of Dispersion of the Power Spectrum, iqr- Interquartile Range, min- Minimum Value, CFREQ- Centroidal Frequency, RHL- Ratio Between Power in High Frequency and Low Frequency, MF- Medium Frequency (4-7Hz), VHF- Very High Frequency (>7Hz), HF- Hight Frequency (>4Hz), LF- Low Frequency (0.15-3.5Hz), trem- Tremor, post- Postural, a- Accelerometer Average Signal, x- Accelerometer Mediolateral Signal, y- Accelerometer Vertical Signal, z- Accelerometer Anteroposterior Signal, Hz- Hertz, Voice Task: c_mean- Mean of the MFCCs Coefficients, log-Energy of the Signal and the First and Second Derivatives of the MFCCs, MFCC- Mel Frequency Cepstral Coefficients, gqc- Glottis Quotient Close, p95- 95th Percentile, shbd- Shimmer, Tapping Task: iqr- Interquartile Range, TapInter- Tap Interval, buttonNoneFreq: Frequency of Tapping Outside the Button, numberTaps- Number of Taps, DriftRight- Right Drift, corXY- Correlation of X and Y Positions, DriftLeft- Left Drift, mad- Median Absolute Deviation, min- Minimum, cv- Coefficient, Sd- Standard Deviation.

### Repetition effects

Next, we evaluated the longitudinal stability of these most reliable features. Using repeated measures analyses of variance, we tested for main effects of diagnosis, repetition (1^st^, 2^nd^, 3^rd^, 4^th,^and 5^th^) and their interaction (Figure 3,4, Table S5, S10-S13). A significant main effect of diagnosis across all time points was observed for 6 out of 15 gait features, 11 out of 15 balance features, 8 out of 12 voice features and 11 out of 12 tapping features. A significant effect of repetition was found for 8 out of 15 gait features, 8 out of 15 balance features, 4 out of 12 voice features 10 out of 12 tapping features. A significant diagnosis-by-repetition interaction effect was identified for 3 out of 15 gait features, 0 out of 15 balance features, 3 out of 12 voice features, and 9 out of 12 tapping features.

**Figure 3.**
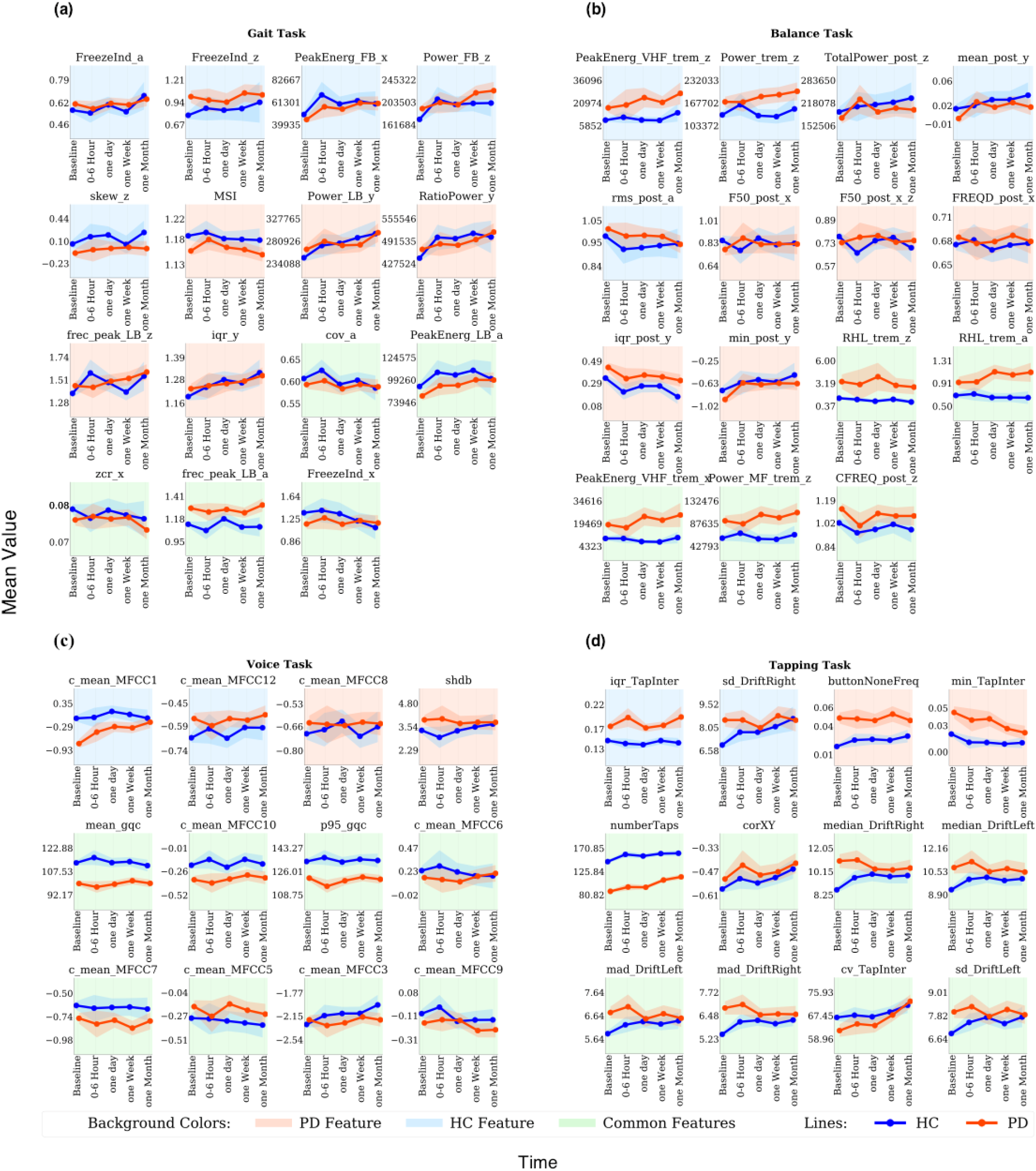
Mean value of the best performing features at baseline and across different time points, calculated for PD and HC separately. **(a)** Gait Task: FreezeInd- Freeze Index, PeakEnerg- Peak of Energy, skew- Skewness, MSI- Mean Stride Interval, RatioPower - Sum of the Power in the Freezing and Locomotor Band, frec_peak- Frequency at the Peak of Energy, iqr- Interquartile Range, cov-Coefficient of Variation, zcr- Zero-Crossing Rate, LB- Locomotor Band, FB- Freezing Band, a- Accelerometer Average Signal, x- Accelerometer Mediolateral Signal, y- Accelerometer Vertical Signal, z- Accelerometer Anteroposterior Signal, **(b)** Balance Task: PeakEnergy - Peak of energy, TotalPower- Energy between .15-3.5 Hz, Power- Energy between 3.5-15Hz, rms- Root Mean Square, F50- Frequency Containing 50% of Total Power, FRQD- Frequency of Dispersion of the Power Spectrum, iqr- Interquartile Range, min- Minimum Value, CFREQ- Centroidal Frequency, RHL- Ratio Between Power in High Frequency and Low Frequency, MF- Medium Frequency (4-7Hz), VHF- Very High Frequency (>7Hz), HF- Hight Frequency (>4Hz), LF- Low Frequency (0.15-3.5Hz), trem- Tremor, post- Postural, a- Accelerometer Average Signal, x- Accelerometer Mediolateral Signal, y- Accelerometer Vertical Signal, z- Accelerometer Anteroposterior Signal, Hz- Hertz, **(c)** Voice task c_mean- Mean of the MFCCs Coefficients, log-Energy of the Signal and the First and Second Derivatives of the MFCCs, MFCC- Mel Frequency Cepstral Coefficients, gqc- Glottis Quotient Close, p95- 95th Percentile, shbd- Shimmer, **(d)** Tapping Task: iqr- Interquartile Range, TapInter- Tap Interval, buttonNoneFreq: Frequency of Tapping Outside the Button, numberTaps- Number of Taps, DriftRight- Right Drift, corXY- Correlation of X and Y Positions, DriftLeft- Left Drift, mad- Median Absolute Deviation, min- Minimum, cv- Coefficient, Sd- Standard Deviation.

**Figure 4.**
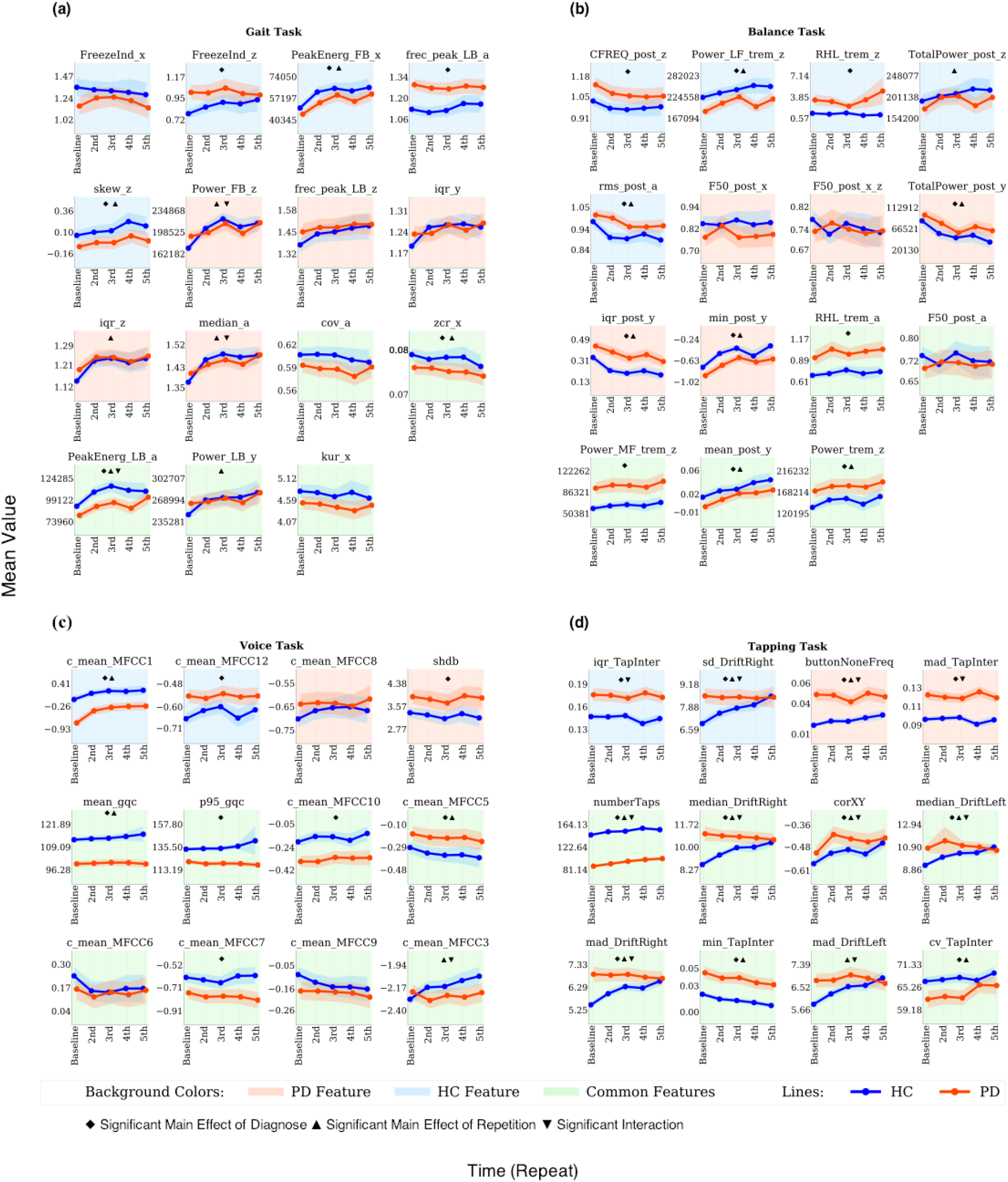
Mean value of the best performing features at baseline and across different time points, calculated for PD and HC separately. **(a)** Gait Task: FreezeInd- Freeze Index, PeakEnerg- Peak of Energy, frec_peak- Frequency at the Peak of Energy, skew- Skewness, iqr- Interquartile Range, cov- Coefficient of Variation, zcr- Zero-Crossing Rate, kur-Kurtosis, LB- Locomotor Band, FB- Freezing Band, a- Accelerometer Average Signal, x- Accelerometer Mediolateral Signal, y- Accelerometer Vertical Signal, z- Accelerometer Anteroposterior Signal, **(b)** Balance Task: PeakEnergy - Peak of Energy, TotalPower- Energy between .15-3.5 Hz, Power- Energy between 3.5-15Hz, rms- Root Mean Square, F50- Frequency Containing 50% of Total Power, FRQD- Frequency of Dispersion of the Power Spectrum, iqr- Interquartile Range, min- Minimum Value, CFREQ- Centroidal Frequency, RHL- Ratio Between Power in High Frequency and Low Frequency, MF- Medium Frequency (4-7Hz), VHF- Very High Frequency (>7Hz), HF- Hight Frequency (>4Hz), LF- Low Frequency (0.15-3.5Hz), trem- Tremor, post- Postural, a- Accelerometer Average Signal, x- Accelerometer Mediolateral Signal, y- Accelerometer Vertical Signal, z- Accelerometer Anteroposterior Signal, Hz- Hertz, **(c)** Voice Task: c_mean- Mean of the MFCCs Coefficients, log-Energy of the Signal and the First and Second Derivatives of the MFCCs, MFCC- Mel Frequency Cepstral Coefficients, shbd- Shimmer, gqc- Glottis Quotient Close, p95- 95th Percentile, **(d)** Tapping Task: Tapping Task: iqr- Interquartile Range, TapInter- Tap Interval, buttonNoneFreq: Frequency of tapping outside the button, numberTaps- Number of Taps, DriftRight- Right Drift, corXY- Correlation of X and Y Positions, DriftLeft- Left Drift, mad- Median absolute deviation, min- Minimum, cv- Coefficient, Sd- Standard Deviation.

### Medication effects

Lastly, we tested which of the most reliable features identified above also display sensitivity to PD medication. For this we compared the conditions reported by the patients as being before PD medication, after PD medication or at best. A significant effect of PD medication was only observed for 2 out of 15 gait features, 1 out of 15 balance features, 2 out of 12 voice features and 1 out of 12 tapping features (Figure S3, Table S5, S10-S13-medication column)

## Discussion

Here we assessed the longitudinal test-retest reliability and stability of DB measures related to gait, balance voice and finger dexterity impairments in PD. We found a wide range of test-retest reliabilities across tasks and features ranging from poor to excellent with highest reliabilities observed for voice followed by the tapping task. Only very few features had medium to large effects sizes for differentiation between PD and HC. For all tasks, a substantial percentage of features displayed significant longitudinal alterations.

Overall, tapping and voice tasks revealed a better performance compared to gait and balance tasks with respect to test-retest reliability and observed effect sizes. Both, balance and gait tasks displayed consistently poor test-retest reliabilities as well as low effect sizes for differentiation between PD and HC questioning their usability for home-based applications. In contrast, best performing voice features displayed fair to excellent test-retest reliabilities across repetitions but also over weeks and months. Most features showed a drop in test-retest reliability with longer periods of time. This may potentially reflect a consequence of the repetition effects and the group-by-repetition interactions observed in the analyses of variance for a substantial proportion of the features.

Despite significant difference at baseline, several features did not differentiate PD and HC when using data from all time points. This effect became most pronounced for the gait task, likely due to its poor test-retest reliability performance. Differential learning, variation in motivation, medication, reduced adherence to task instructions, and other physical and environmental parameters may contribute to this loss of differentiation [2][9,11]. Whilst a clear differentiation of motivation vs learning effects on the often abstract DB features is difficult in an observational study design, a possible way to provide inference on this issue is to compare the direction of alterations in PD and HC. Assuming that alterations in PD relative to HC reflect impairment, movement of a feature state towards PD is likely to reflect worsening either due to reduced motivation, disease progression or other similar factors. In contrast, movements towards HC is likely to reflect improvement and is therewith compatible with a learning effect. We find a mixture of both effects for most tasks suggesting the presence of both aspects in DB longitudinal data. These observations are also in line with previous studies showing that training may reduce motor impairment in PD [34– 36]. In particular, for the tapping task the difference between PD and HC disappears for several features which is primarily due to a shift in performance in HC. These findings may point to a differential change in motivation across groups. Whilst differential learning has been previously reported [34,37,38], the differential change in motivation is an important novel aspect to consider when comparing DB measures between PD patients and HC. Understanding the sources leading to this variability of DB measures over time is a vital and open-question which needs to be systematically addressed to enable their application for specific clinical questions.

Most patients with PD take dopaminergic medication to alleviate their motor functions. However, the responsiveness to PD medication highly varies between patients. Besides good reliability and the ability to differentiate PD and HC, another important and desired quality of an effective DB is therefore to monitor PD medication response. Among the most reliable features from each task, only very few displayed significant but weak sensitivity to different medication conditions. One possible reason for this poor performance of DB measures in our study as compared to some previous reports [39], might be the self-reported nature of the medication status in the m-Power dataset likely introducing some noise variation (i.e. different drugs and differences in time after administration). Nonetheless, our findings point to the need for further optimization of DB measures to increase their sensitivity to PD medication effects.

The self-administered design of the m-Power dataset is also the major limitation of our study. In such an uncontrolled setting, accuracy in reporting the diagnosis and demographics, defining the medication status but also correct understanding and compliance with the instructions may all have introduced variation into the study measures. The reported ballpark estimates for test-retest reliability and ability of the respective measures to differentiate between PD and HC therefore need to be carefully considered when interpreting our results. Nonetheless, our findings clearly demonstrate the need for further optimization of DB tasks as well for introducing careful monitoring and quality control procedures to enable integration of DB measures into clinically relevant applications.

## Data Availability

The m-Power dataset used for this article is available upon registration from Synapse at: https://www.synapse.org/#!Synapse:syn4993293/

## Data availability

The m-Power dataset used for this article is available upon registration from Synapse at: https://www.synapse.org/#!Synapse:syn4993293.

## Acknowledgements

This study was supported by the Human Brain Project, funded from the European Union’s Horizon 2020 Framework Programme for Research and Innovation under the Specific Grant Agreement No. 785907 (Human Brain Project SGA2).

The data used in this study were contributed by users of the Parkinson m-Power mobile application as part of the m-Power study developed by Sage Bionetworks and described in Synapse [doi:10.7303/syn4993293].

## Author contribution

MSF performed analyses and wrote manuscript. MG performed feature extraction. MSF, JD and MG contributed to study design and writing the manuscript. SBE and JD designed the overall study and contributed to interpretation of the results. All authors reviewed and commented on the manuscript.

## References

1. Jankovic J. Parkinson’s disease: clinical features and diagnosis. J Neurol Neurosurg Psychiatry 2008 Apr;79(4):368–376. PMID: 18344392

2. Prince J, Arora S, de Vos M. Big data in Parkinson’s disease: using smartphones to remotely detect longitudinal disease phenotypes. Physiol Meas 2018 Apr 26;39(4):044005. PMID: 29516871

3. Insel TR. Digital phenotyping: technology for a new science of behavior. JAMA 2017 Oct 3;318(13):1215–1216. PMID: 28973224

4. Mahadevan N, Demanuele C, Zhang H, Volfson D, Ho B, Erb MK, et al. Development of digital biomarkers for resting tremor and bradykinesia using a wrist-worn wearable device. npj Digital Med 2020 Jan 15;3:5. PMID: 31970290

5. Lipsmeier F, Taylor KI, Kilchenmann T, Wolf D, Scotland A, Schjodt-Eriksen J, et al. Evaluation of smartphone-based testing to generate exploratory outcome measures in a phase 1 Parkinson’s disease clinical trial. Mov Disord 2018 Apr 27;33(8):1287–1297. PMID: 29701258

6. Shah VV, McNames J, Mancini M, Carlson-Kuhta P, Nutt JG, El-Gohary M, et al. Digital biomarkers of mobility in parkinson’s disease during daily living. J Parkinsons Dis 2020;10(3):1099–1111. PMID: 32417795

7. Schlachetzki JCM, Barth J, Marxreiter F, Gossler J, Kohl Z, Reinfelder S, et al. Wearable sensors objectively measure gait parameters in Parkinson’s disease. PLoS One 2017 Oct 11;12(10):e0183989. PMID: 29020012

8. Tracy JM, Özkanca Y, Atkins DC, Hosseini Ghomi R. Investigating voice as a biomarker: Deep phenotyping methods for early detection of Parkinson’s disease. J Biomed Inform 2020 Apr;104:103362. PMID: 31866434

9. Espay AJ, Bonato P, Nahab FB, Maetzler W, Dean JM, Klucken J, et al. Technology in Parkinson’s disease: Challenges and opportunities. Mov Disord 2016 Apr 29;31(9):1272– 1282. PMID: 27125836

10. Moore ST, Yungher DA, Morris TR, Dilda V, MacDougall HG, Shine JM, et al. Autonomous identification of freezing of gait in Parkinson’s disease from lower-body segmental accelerometry. J Neuroeng Rehabil 2013 Feb 13;10:19. PMID: 23405951

11. Fisher JM, Hammerla NY, Rochester L, Andras P, Walker RW. Body-Worn Sensors in Parkinson’s Disease: Evaluating Their Acceptability to Patients. Telemed J E Health 2016 Jan;22(1):63–69. PMID: 26186307

12. Rahlf AL, Petersen E, Rehwinkel D, Zech A, Hamacher D. Validity and Reliability of an Inertial Sensor-Based Knee Proprioception Test in Younger vs. Older Adults. Front Sports Act Living 2019 Sep 18;1.

13. Orlowski K, Eckardt F, Herold F, Aye N, Edelmann-Nusser J, Witte K. Examination of the reliability of an inertial sensor-based gait analysis system. Biomed Tech (Berl) 2017 Nov 27;62(6):615–622. PMID: 28099115

14. Hasegawa N, Shah VV, Carlson-Kuhta P, Nutt JG, Horak FB, Mancini M. How to Select Balance Measures Sensitive to Parkinson’s Disease from Body-Worn Inertial Sensors-Separating the Trees from the Forest. Sensors (Basel) 2019 Jul 28;19(15). PMID: 31357742

15. Skodda S, Grönheit W, Mancinelli N, Schlegel U. Progression of voice and speech impairment in the course of Parkinson’s disease: a longitudinal study. Parkinsons Dis 2013 Dec 10;2013:389195. PMID: 24386590

16. Aghanavesi S, Nyholm D, Senek M, Bergquist F, Memedi M. A smartphone-based system to quantify dexterity in Parkinson’s disease patients. Informatics in Medicine Unlocked 2017;9:11–17.

17. Wissel BD, Mitsi G, Dwivedi AK, Papapetropoulos S, Larkin S, López Castellanos JR, et al. Tablet-Based Application for Objective Measurement of Motor Fluctuations in Parkinson Disease. Digit Biomark 2018 Jan 9;1(2):126–135.

18. Bot BM, Suver C, Neto EC, Kellen M, Klein A, Bare C, et al. The mPower study, Parkinson disease mobile data collected using ResearchKit. Sci Data 2016 Mar 3;3:160011. PMID: 26938265

19. Mirelman A, Heman T, Yasinovsky K, Thaler A, Gurevich T, Marder K, et al. Fall risk and gait in Parkinson’s disease: the role of the LRRK2 G2019S mutation. Mov Disord 2013 Oct 7;28(12):1683–1690. PMID: 24123150

20. Blin O, Ferrandez AM, Serratrice G. Quantitative analysis of gait in Parkinson patients: increased variability of stride length. J Neurol Sci 1990 Aug;98(1):91–97. PMID: 2230833

21. Hausdorff JM. Gait dynamics in Parkinson’s disease: common and distinct behavior among stride length, gait variability, and fractal-like scaling. Chaos 2009 Jun;19(2):026113. PMID: 19566273

22. Miller Koop M, Ozinga SJ, Rosenfeldt AB, Alberts JL. Quantifying turning behavior and gait in Parkinson’s disease using mobile technology. IBRO Rep 2018 Dec;5:10–16. PMID: 30135951

23. Martinez-Mendez R, Sekine M, Tamura T. Postural sway parameters using a triaxial accelerometer: comparing elderly and young healthy adults. Comput Methods Biomech Biomed Engin 2012;15(9):899–910. PMID: 21547782

24. Prieto TE, Myklebust JB, Hoffmann RG, Lovett EG, Myklebust BM. Measures of postural steadiness: differences between healthy young and elderly adults. IEEE Trans Biomed Eng 1996 Sep;43(9):956–966. PMID: 9214811

25. Palakurthi B, Burugupally SP. Postural instability in parkinson’s disease: A review. Brain Sci 2019 Sep 18;9(9). PMID: 31540441

26. Pittman B, Ghomi RH, Si D. Parkinson’s Disease Classification of mPower Walking Activity Participants. Annu Int Conf IEEE Eng Med Biol Soc 2018 Jul;2018:4253–4256. PMID: 30441293

27. Theory B, Algorithm. Implementing Positioning Algorithms Using Accelerometers.

28. Palmerini L, Rocchi L, Mellone S, Valzania F, Chiari L. Feature selection for accelerometer-based posture analysis in Parkinson’s disease. IEEE Trans Inf Technol Biomed 2011 May;15(3):481–490. PMID: 21349795

29. Little MA, McSharry PE, Roberts SJ, Costello DAE, Moroz IM. Exploiting nonlinear recurrence and fractal scaling properties for voice disorder detection. Biomed Eng Online 2007 Jun 26;6:23. PMID: 17594480

30. Chiaramonte R, Bonfiglio M. Acoustic analysis of voice in Parkinson’s disease: a systematic review of voice disability and meta-analysis of studies. Rev Neurol 2020 Jun 1;70(11):393– 405. PMID: 32436206

31. Rao G, Fisch L, Srinivasan S, D’Amico F, Okada T, Eaton C, et al. Does this patient have Parkinson disease? JAMA 2003 Jan 15;289(3):347–353. PMID: 12525236

32. Jobbágy A, Harcos P, Karoly R, Fazekas G. Analysis of finger-tapping movement. J Neurosci Methods 2005 Jan 30;141(1):29–39. PMID: 15585286

33. Cicchetti DV. Guidelines, criteria, and rules of thumb for evaluating normed and standardized assessment instruments in psychology. Psychol Assess 1994;6(4):284–290.

34. Olson M, Lockhart TE, Lieberman A. Motor learning deficits in parkinson’s disease (PD) and their effect on training response in gait and balance: A narrative review. Front Neurol 2019 Feb 7;10:62. PMID: 30792688

35. Steib S, Wanner P, Adler W, Winkler J, Klucken J, Pfeifer K. A single bout of aerobic exercise improves motor skill consolidation in parkinson’s disease. Front Aging Neurosci 2018 Oct 22;10:328. PMID: 30405397

36. Bryant M. Effect of Resistance Exercise on Tremor and Hand Dexterity of Parkinson’s Disease.

37. Krebs HI, Hogan N, Hening W, Adamovich SV, Poizner H. Procedural motor learning in Parkinson’s disease. Exp Brain Res 2001 Dec;141(4):425–437. PMID: 11810137

38. Sehm B, Taubert M, Conde V, Weise D, Classen J, Dukart J, et al. Structural brain plasticity in Parkinson’s disease induced by balance training. Neurobiol Aging 2014 Jan;35(1):232–239. PMID: 23916062

39. Zhan A, Little MA, Harris DA, Abiola SO, Dorsey ER, Saria S, et al. High Frequency Remote Monitoring of Parkinson’s Disease via Smartphone: Platform Overview and Medication Response Detection. arXiv 2016 Jan 5;

