## supplement for "Exploring test retest reliability and longitudinal stability of digital biomarkers for Parkinson’s disease in the m-Power dataset"

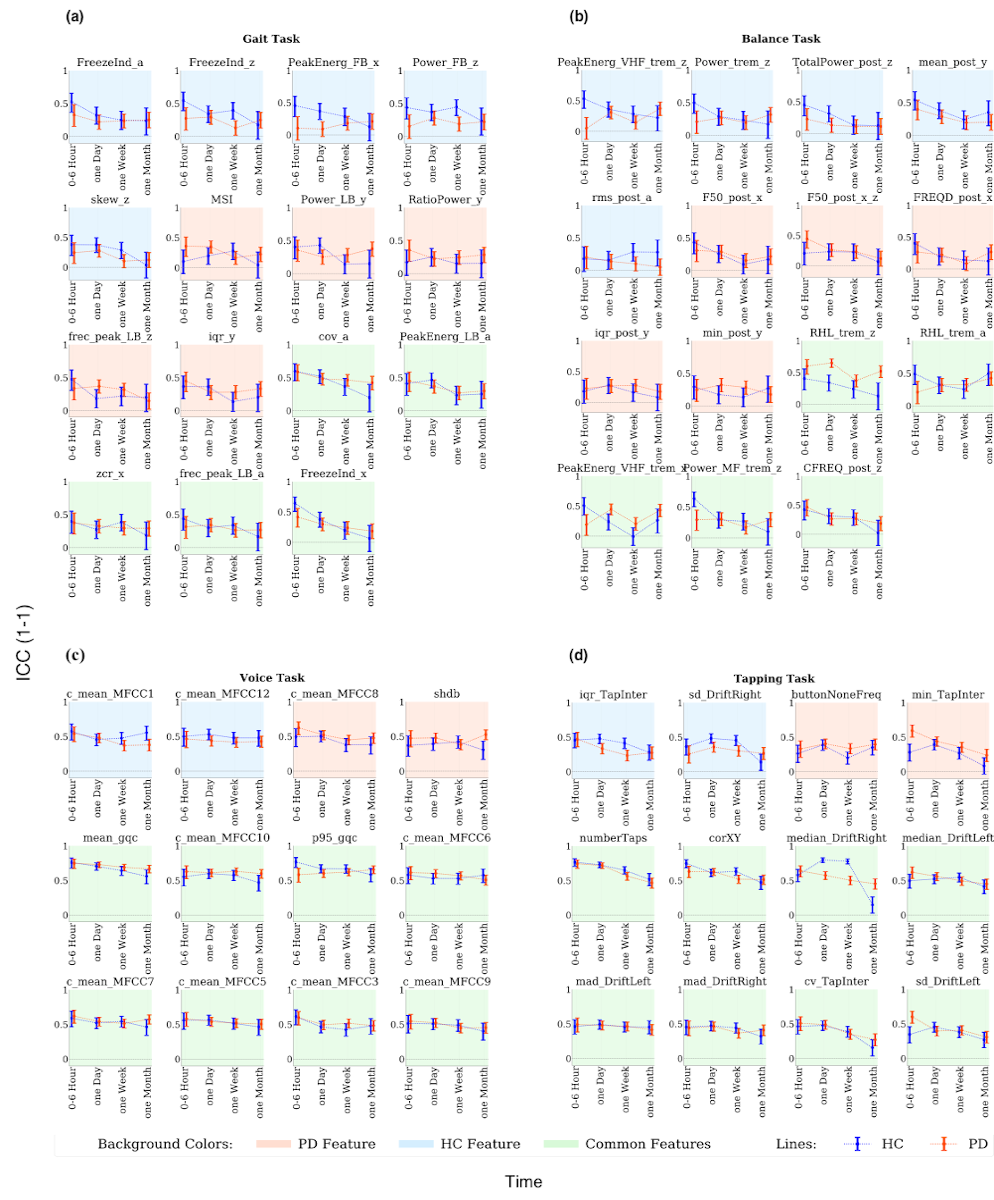

**Figure S1** Test-retest reliability for the best performing features. Intraclass Correlation Coefficient ICCs (1-1) Value with a 95% confidence interval across different time points vs baseline. **(a)** Gait Task: FreezeInd- Freeze Index, PeakEnerg- Peak of Energy, skew- Skewness, MSI- Mean Stride Interval, RatioPower - Sum of the Power in the Freezing and Locomotor Band, frec_peak- Frequency at the Peak of Energy, iqr- Interquartile Range, cov- Coefficient of Variation, zcr- Zero-Crossing Rate, LB- Locomotor Band, FB- Freezing Band, a- Accelerometer Average Signal,x- Accelerometer Mediolateral Signal, y- Accelerometer Vertical Signal, z- Accelerometer Anteroposterior Signal, **(b)** Balance Task: PeakEnergy- Peak of energy, TotalPower- Energy between .15-3.5 Hz, Power- Energy between 3.5-15Hz, rms- Root Mean Square, F50- Frequency Containing 50% of Total Power, FRQD- Frequency of Dispersion of the Power Spectrum, iqr- Interquartile Range, min- Minimum Value, CFREQ- Centroidal Frequency, RHL- Ratio Between Power in High Frequency and Low Frequency, MF- Medium Frequency (4-7Hz), VHF- Very High Frequency (>7Hz) , HF- Hight Frequency (>4Hz), LF- Low Frequency (0.15-3.5Hz), trem- Tremor, post- Postural, a- Accelerometer Average Signal, x- Accelerometer Mediolateral Signal, y- Accelerometer Vertical Signal, z- Accelerometer Anteroposterior Signal, Hz- Hertz, **(c)** Voice task : c_mean_MFCC1–12- Mean Value of Mel Frequency Cepstral Coefficients 1-12, gqc- Glottis Quotient Close, p95- 95th Percentile, **(d)** Tapping Task: iqr- Interquartile Range, TapInter- Tap Interval, buttonNoneFreq: Frequency of Tapping outside the Button, numberTaps- Number of Taps, DriftRight- Right Drift, corXY- Correlation of X and Y Positions, DriftLeft- Left Drift, mad- Median Absolute Deviation, min- Minimum, cv- Coefficient, Sd- Standard Deviation.

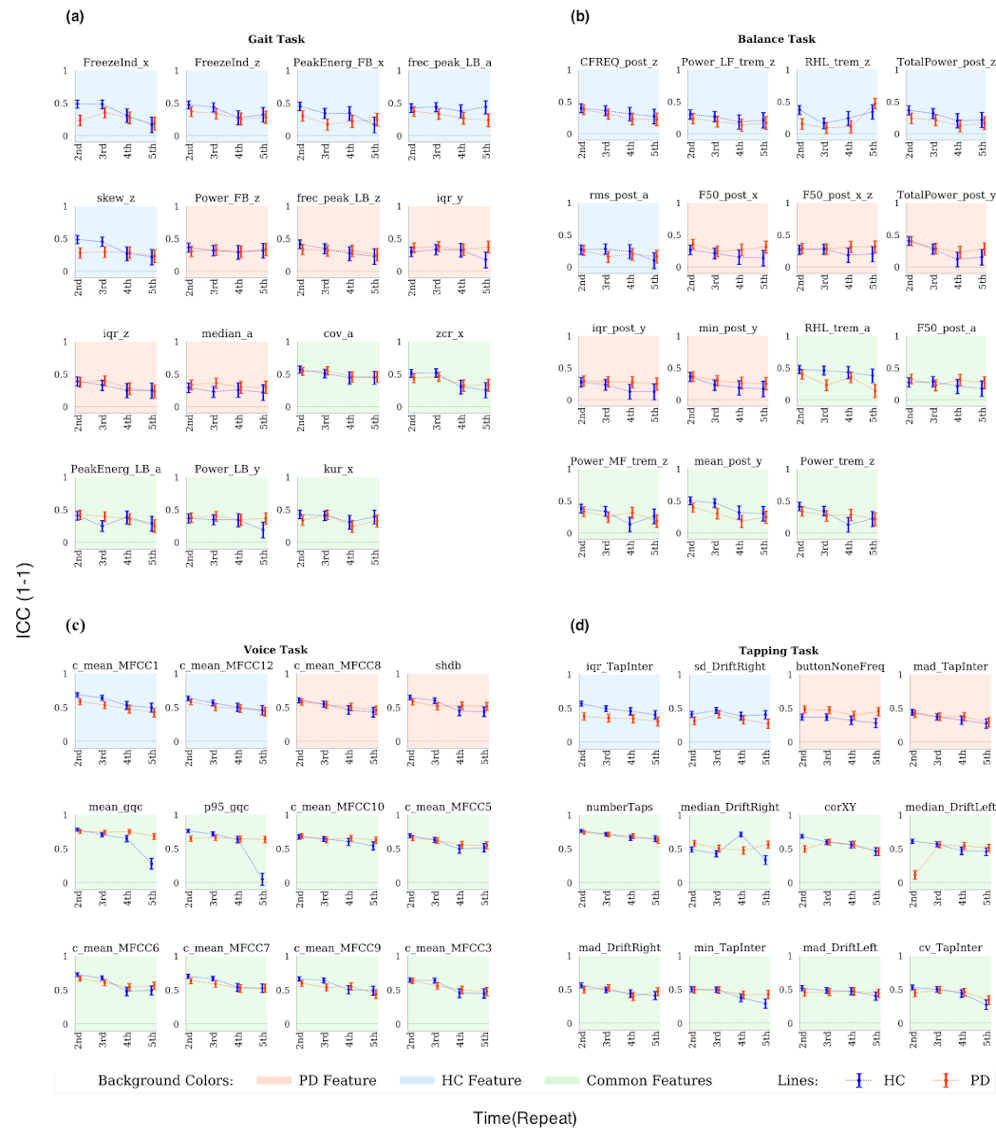

**Figure S2** Test-retest reliability for the best performing features. Intraclass Correlation Coefficient ICCs (1-1) Value with a 95% confidence interval across different repetition vs baseline. **(a)** Walking Task: FreezeInd- Freeze Index, PeakEnerg- Peak of Energy, frec_peak- Frequency at the Peak of Energy, skew- Skewness, iqr- Interquartile Range, cov- Coefficient of Variation, zcr- Zero-Crossing Rate, kur-Kurtosis, LB- Locomotor Band, FB- Freezing Band, a- Accelerometer Average Signal, x- Accelerometer Mediolateral Signal, y- Accelerometer Vertical Signal, z- Accelerometer Anteroposterior Signal, **(b)** Balance Task: PeakEnergy - Peak of energy, TotalPower- Energy between .15-3.5 Hz, Power- Energy between 3.5-15Hz, rms- Root Mean Square, F50- Frequency Containing 50% of Total Power, FRQD- Frequency of Dispersion of the Power Spectrum, iqr- Interquartile Range, min- Minimum Value, CFREQ- Centroidal Frequency, RHL- Ratio Between Power in High Frequency and Low Frequency, MF- Medium Frequency (4-7Hz), VHF- Very High Frequency (>7Hz) , HF- Hight Frequency (>4Hz), LF- Low Frequency (0.15-3.5Hz), trem- Tremor, post- Postural, a- Accelerometer Average Signal, x- Accelerometer Mediolateral Signal, y- Accelerometer Vertical Signal, z- Accelerometer Anteroposterior Signal, Hz- hertz, **(c)** Voice Task: c_mean_MFCC1–12- Mean Value of Mel Frequency Cepstral Coefficients 1-12, shbd- Shimmer, gqc- Glottis Quotient Close, p95- 95th Percentile, **(d)** Tapping Task: Tapping Task: iqr- Interquartile Range, TapInter- Tap Interval, buttonNoneFreq: Frequency of Tapping Outside the Button, numberTaps- Number of Taps, DriftRight- Right Drift, corXY- Correlation of X and Y Positions, DriftLeft- Left Drift, mad- Median Absolute Deviation, min- Minimum, cv- Coefficient, Sd- Standard Deviation.

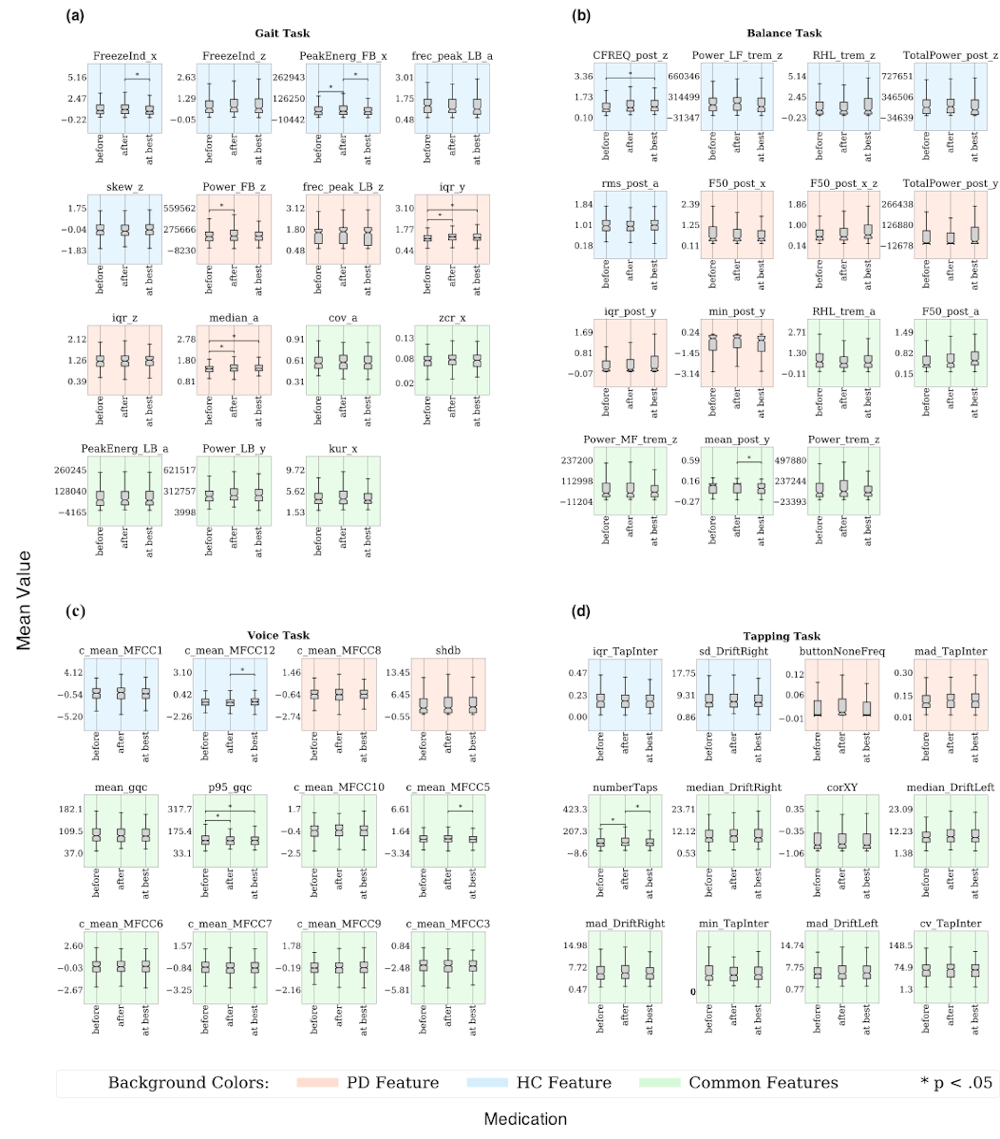

**Figure S3** Mean Value of the best performing features at different medication conditions. **(a)** Gait Task: FreezeInd- Freeze Index, PeakEnerg- Peak of Energy, frec_peak- Frequency at the Peak of Energy, skew- Skewness, iqr- Interquartile Range, cov- Coefficient of Variation, zcr- Zero-Crossing Rate, kur-Kurtosis, LB- Locomotor Band, FB- Freezing Band, a- Accelerometer Average Signal, x- Accelerometer Mediolateral Signal, y- Accelerometer Vertical Signal, z- Accelerometer Anteroposterior Signal, **(b)** Balance Task: PeakEnergy - Peak of energy, TotalPower- Energy between .15-3.5 Hz, Power- Energy between 3.5-15Hz, rms- Root Mean Square, F50- Frequency Containing 50% of Total Power, FRQD- Frequency of Dispersion of the Power Spectrum, iqr- Interquartile Range, min- Minimum Value, CFREQ- Centroidal Frequency, RHL- Ratio Between Power in High Frequency and Low Frequency, MF- Medium Frequency (4-7Hz), VHF- Very High Frequency (>7Hz) , HF- Hight Frequency (>4Hz), LF- Low Frequency (0.15-3.5Hz), trem- Tremor, post- Postural, a- Accelerometer Average Signal, x- Accelerometer Mediolateral Signal, y- Accelerometer Vertical Signal, z- Accelerometer Anteroposterior Signal, Hz- hertz, **(c)** Voice Task: c_mean_MFCC1–12- Mean Value of Mel Frequency Cepstral Coefficients 1-12, shbd- Shimmer, gqc- Glottis Quotient Close, p95- 95th Percentile, **(d)** Tapping Task: Tapping Task: iqr- Interquartile Range, TapInter- Tap Interval, buttonNoneFreq: Frequency of tapping outside the button, numberTaps- Number of Taps, DriftRight- Right Drift, corXY- Correlation of X and Y Positions, DriftLeft- Left Drift, mad- Median Absolute Deviation, min- Minimum, cv- Coefficient, Sd- Standard Deviation.

**Table S1** Gait Features.

| Feature acronym | Units | Feature description | Signal (acronym) |
| --- | --- | --- | --- |
| numSteps |  | Number of steps during the 10 seconds gait signal. |  |
| MSI | s | Mean Stride Interval, calculated as the duration of a stride averaged over all strides[1,2]. |  |
| StrideVar | % | Stride Variability, calculated as the standard deviation divided by the mean stride of the stride interval. Measures consistency and stability[1,2]. |  |
| mean | ms^-2 | Mean value of the observations[3,4]. | Mediolateral, vertical, anteroposterior, and average acceleration (x, y, z, a)  Mediolateral, vertical, anteroposterior, and average postural acceleration (post_x, post _y, post _z, post_a)  velocity (vel)  acceleration along path (acc) |
| min | ms^-2 | Minimum value of the observations. |  |
| max | ms^-2 | Maximum value of the observations. |  |
| median | ms^-2 | Median. Middle value among a dataset[3,4]. |  |
| sd | ms^-2 | Standard deviation, calculated as the sum of squares differences between the individual values and the mean. Measures variability[3,4]. |  |
| var | (ms^2)^2 | Variance, calculated as the square of the standard deviation. Measures variability. |  |
| range | ms^-2 | Range of the observations. |  |
| iqr | ms^-2 | Interquartile range, calculated as the difference between 75^th^ and 25^th^ percentiles. Measures dispersion[3,4]. |  |
| rms | ms^-2 | Root mean square of the observations. |  |
| cov |  | Coefficient of variation, calculated as the standard deviation of the signal divided by the mean. |  |
| skew |  | Skewness. Describes the asymmetry of a signal. A negative value indicates that the distribution is concentrated on the right, while a positive one is concentrated in the left[2–4]. |  |
| kur |  | Kurtosis. Measures if data is heavy or light-tailed to a normal distribution[2,3]. |  |
| zcr |  | Zero-crossing rate. Rates sign-changes along a signal[4]. |  |
| ApEn |  | Entropy. Measures uncertainty, ranging from 0-1 where 0 indicates randomness and 1 maximum regularity[2,4]. |  |
| PeakEnerg_LB | psd | Peak of energy in the locomotor band (0.5-3 Hz)[1,5]. |  |
| frec_peak_LB | Hz | Frequency at the peak of energy in the locomotor band (0.5-3 Hz)[5]. |  |
| Power_LB | psd | Power of the locomotor band (0.5-3 Hz)[5]. |  |
| PeakEnerg_FB | psd | Peak of energy in the freezing band (3-8Hz)[6]. |  |
| frec_peak_FB | Hz | Frequency at the peak of energy in the freezing band (3-8 Hz). |  |
| Power_FB | psd | Power in the freezing band (3-8 Hz). |  |
| FreezeInd |  | Freeze Index. Calculated as the ratio between the power in the freezing band (3-8 Hz) and the power in the locomotor band (0.5-3 Hz)[6]. |  |
| RatioPower | psd | Sum of the power in the freezing (3-8 Hz) and locomotor band (3-8 Hz)[7] |  |
| ar |  | Coefficient of a 1^st^ order autoregressive model. An autoregressive model forecasts when there is some correlation between current values and their preceding ones. |  |
| COEFCEPS_(1-20) | mel | 20 Mel Frequency Cepstral Coefficients. Represent the short-term power spectrum[6] |  |

**Table S2** Balance Features.

| Acronym | units | Description | Signal (acronym) |
| --- | --- | --- | --- |
| mean | ms^-2 | Mean value of the observations. | Mediolateral, vertical, anteroposterior, and average tremor acceleration (trem_x, trem_y, trem_z, trem_a)  Mediolateral, vertical, anteroposterior, and average postural acceleration (post_x, post _y, post _z, post_a) |
| min | ms^-2 | Minimum value of the observations. |  |
| max | ms^-2 | Maximum value of the observations. |  |
| median | ms^-2 | Median value of the observations. |  |
| sd | ms^-2 | Standard deviation, calculated as the sum of squares differences between the individual values and the mean. Measures variability. |  |
| var | mg^2 | Variance, calculated as the square of the standard deviation. Measures variability. |  |
| range | ms^-2 | Range of the observations. |  |
| iqr | ms^-2 | Interquartile range, calculated as the difference between 75^th^ and 25^th^ percentiles. Measures dispersion. |  |
| rms | ms^-2 | Root mean square of the observations. |  |
| cov | ms^-2 | Coefficient of variation, calculated as the standard deviation of the signal divided by the mean. |  |
| skew |  | Skewness. Describes the asymmetry of a signal. A negative value indicates that the distribution is concentrated on the right, while a positive one is concentrated in the left. |  |
| kur |  | Kurtosis. Measures if data is heavy or light-tailed to a normal distribution. |  |
| zcr |  | Zero-crossing rate. Rates sign-changes along a signal. |  |
| ApEn |  | Entropy. Measures uncertainty, ranging from 0-1 where 0 indicates randomness and 1 maximum regularity. |  |
| Power_MF | psd | Power of the medium frequency band 4-7Hz [8]. | Mediolateral, vertical, anteroposterior, and average tremor acceleration (trem_x, trem_y, trem_z, trem_a) |
| PeakEnerg_VHF | psd | Peak of energy in the very high frequency band (>7Hz) |  |
| frec_peak_HF | Hz | Frequency at the peak of energy in high frequency band (>4Hz)[8]. |  |
| Power | psd | Power between 3.5-15Hz |  |
| Power_LF | psd | Power in the low frequency band (0.15-3.5Hz) |  |
| RHL |  | Ratio between the power between 3.5-15Hz and power between 0.15-3.5Hz[8]. |  |
| CFREQ | Hz | Centroidal frequency for postural measures. Also known as zero-crossing frequency[8–10]. | Mediolateral, vertical, anteroposterior, and average postural acceleration (post_x, post _y, post _z, post_a)  Mediolateral-anteroposterior average postural acceleration (post_x_z) |
| FREQD | Hz | Frequency of dispersion of the power spectrum for postural measures[8–10]. |  |
| jerk | m^2/s^2 | Average jerk. Measures vibration as the rate of change in acceleration. Calculated as the derivative of acceleration with respect to time[8,9]. | Mediolateral, vertical, anteroposterior, and average postural acceleration (post_x, post _y, post _z, post_a)  Mediolateral-anteroposterior average postural acceleration (post_x_z) |
| TotalPower | pwd | Energy between 0.15-3.5Hz for postural measures[9]. |  |
| F50 | Hz | Frequency containing 50% of the total power for postural measures[8,9]. |  |
| F95 | Hz | Frequency containing 95% of the total power for postural measures[8,9]. |  |
| MDIST | mm | Represents the average distance from the center to each AP and ML points[8,10]. | Mediolateral, anteroposterior and average of mediolateral and anteroposterior distance (dist_x, dist_z,dist_x_z) |
| RDIST | mm | Root Mean Square distance from the mean center[9,10]. |  |
| TOTEX | mm | Total excursions is the total length of the path. Calculated as the sum of distances between consecutive points[10]. |  |
| MVELO | mm/s | Mean velocity is the average velocity of the center path, calculated as the TOTEX divided by the time[8,10]. |  |
| MFREQ | mm/s | The mean frequency is the rotational frequency with a radius equal to the mean distance[9,10]. |  |
| AREA_CC | mm^2 | The 95% confidence circle area is the area of a circle enclosing all points in the AP-ML plane with 95% confidence[9,10]. | average of mediolateral and anteroposterior distance  (dist_x_z) |
| AREA_CE | mm^2 | The 95% confidence ellipse area is the area of an ellipse enclosing all points in the AP-ML plane with 95% confidence[8–10]. |  |
| AREA_SW | mm^2/s | Sway area calculated as the area enclosing the acceleration path[8–10]. |  |
| FD |  | The fractal dimension indicates the degree to which a curve fills the enclosed metric space[9,10]. |  |
| FD_CC |  | Fractal dimension based on the 95% confidence circle area[9,10]. |  |
| FD_CE |  | Fractal dimension based on the 95% confidence ellipse area[9,10]. |  |

**Table S3** Voice Features

| acronym | Units | description | Signal (acronym) |
| --- | --- | --- | --- |
| amp | psd^0.5 | Average amplitude[11]. |  |
| shim | % | Absolute shimmer[4,11,12]. |  |
| shdb | db | Shimmer in logarithmic domain[11]. |  |
| apq3 | % | 3 point amplitude perturbation quotient in percentage[11]. |  |
| apq5 | % | 5 point amplitude perturbation quotient in percentage[11]. |  |
| fm | Hz | Frequency modulation[11]. |  |
| hnr_mean | db | Mean of the harmonic to noise ratio, which indicates the amount of noise[4,11,12]. |  |
| hnr_std | db | Standard deviation of the harmonic to noise ratio[11]. |  |
| rpde |  | Recurrence period density entropy. Characterizes the deviation from signal periodicity[4,11]. |  |
| DFA |  | Detrended Fluctuation Analysis, which describes turbulent noise[4,11]. |  |
| mean |  | Mean value[4,11]. | fundamental frequency (f0), amplitude (amp), Teager Kaiser Energy Operator of the fundamental frequency (tkeo), open quotient (oq), glottis quotient open (gqo),  glottis quotient closed (gqc) |
| sd |  | Standard deviation[4,11]. |  |
| jitt |  | Absolute jitter[4,11]. | fundamental frequency (f0),  period (T) |
| jitta |  | Relative or local jitter[11]. |  |
| rap |  | Relative average perturbation[11]. |  |
| ppq5 | St | Perturbation quotient using 5 point (cycles)[11]. |  |
| range |  | Range[11]. | Teager Kaiser Energy Operator of the fundamental frequency (tkeo), |
| p25 | Hz^2 | 25^th^ percentile of the Teager-Kaiser Energy Operator[11]. |  |
| p75 | Hz^2 | 75^th^ percentile of the Teager-Kaiser Energy Operator[11]. |  |
| ApEn |  | Pitch Period Entropy. Quantifies the impaired control of stable pitch during a sustained phonation[4,11]. |  |
| p5 | Hz^2 (teko) | 5^th^ percentile[11]. | Teager Kaiser Energy Operator of the fundamental frequency (tkeo), open quotient (oq), glottis quotient open (gqo),  glottis quotient closed (gqc) |
| p95 | Hz^2 | 95^th^ percentile[11]. |  |
| c_mean | db, mel, 1st and 2nd derivative of mel | Mean of the MFCCs coefficients, log-energy of the signal and the first and second derivatives of the MFCCs[4,11,12]. | log energy (log),  0^th^ order cepstral coefficient (0th),  1-12^th^ Mel Frequency Cepstral Coefficients (MFCC_(1-12),  1-14^th^ deltas (d_(1-14)),  1-14^th^ delta-delta (dd_(1-14)) |
| c_std | db, mel, 1st and 2nd derivative of mel | Standard deviation of the MFCCs coefficients, log-energy of the signal and the first and second derivatives of the MFCCs[11,12]. |  |

**Table S4** Tapping Features.

| acronym | units | description | Signal (acronym) |
| --- | --- | --- | --- |
| nTaps |  | Number of taps[13,14]. |  |
| buttonNone |  | Frequency of tapping outside the button [13,14]. |  |
| corXY |  | Correlation between X and Y position of tap on screen coordinates [13,14]. |  |
| mean | s | Mean value of the observations[4,13,14]. | Intertap interval (TapInter), Leftdrift (DriftLeft), Right drift (DriftRight) |
| min | s | Minimum value of the observations[4,13,14]. |  |
| max | s | Maximum value of the observations[4,13,14]. |  |
| median | s | Median value of the observations[4,13,14]. |  |
| mad | s | Median absolute deviation[13,14]. |  |
| sd | s | Standard deviation[4,13,14]. |  |
| range | s | Range of the observations[4,13,14]. |  |
| iqr | s | Interquartile range [13,14]. |  |
| cv |  | Coefficient of variation [4,13,14]. |  |
| skew |  | Skewness[13,14]. |  |
| kur |  | Kurtosis[13,14]. |  |
| tkeo |  | Teager-Kaiser Energy Operator. Measures energy variation[4,13,14]. | Intertap interval (TapInter) |
| dfa |  | Detrended Fluctuation Analysis. Measures changes in the signal[4,13,14]. |  |
| ar1 |  | Coefficient of an autoregressive model at lag 1. Indicates associations between intertap intervals[4,13,14]. |  |
| ar2 |  | Coefficient of an autoregressive model at lag 2. Indicates associations between intertap intervals[4,13,14]. |  |
| fatigue10 |  | Increase in the mean intertap interval from the first 10% to the last 10% taps [4,13,14]. |  |
| fatigue25 |  | Increase in the mean intertap interval from the first 25% to the last 25% taps[4,13,14]. |  |
| fatigue50 |  | Increase in the mean intertap interval from the first 50% to the last 50 % taps[4,13,14]. |  |

**Table S5** Results from an analysis of ANOVA for repeated measurements on the best performing features.

| Features | Diagnosis (PD, HC) | | Repetition | | Diagnosis × Repetition | | Medication |
| --- | --- | --- | --- | --- | --- | --- | --- |
|  | F | *P* | F | *P* | F | *P* | n=188 |
| Gait Task | | | | | | | |
| frec_peak_LB_a | 23.202 | <0.001 | 0.767 | 0.547 | 0.276 | 0.894 | 0.358 |
| FreezeInd_z | 3.877 | 0.049 | 0.665 | 0.616 | 1.012 | 0.4 | 0.422 |
| Power_FB_z | 0.144 | 0.704 | 10.235 | <0.001 | 4.719 | <0.001 | 0.08 |
| PeakEnerg_LB_a | 10.268 | 0.001 | 9.974 | <0.001 | 3.02 | 0.017 | 0.376 |
| iqr_z | 0.109 | 0.742 | 6.849 | <0.001 | 1.102 | 0.354 | 0.806 |
| skew_z | 9.226 | 0.002 | 2.749 | 0.027 | 0.267 | 0.899 | 0.192 |
| median_a | 1.003 | 0.317 | 13.362 | <0.001 | 2.814 | 0.024 | 0.058 |
| FreezeInd_x | 2.681 | 0.102 | 0.954 | 0.432 | 0.406 | 0.804 | 0.151 |
| zcr_x | 4.987 | 0.026 | 4.387 | 0.002 | 0.827 | 0.508 | 0.58 |
| Power_LB_y | 0.256 | 0.613 | 2.985 | 0.018 | 0.792 | 0.53 | 0.156 |
| iqr_y | 0 | 1.0 | 2.28 | 0.059 | 1.223 | 0.299 | 0.01 |
| kur_x | 2.438 | 0.119 | 0.418 | 0.796 | 1.969 | 0.097 | 0.375 |
| frec_peak_LB_z | 1.013 | 0.315 | 2.198 | 0.067 | 0.705 | 0.589 | 0.449 |
| PeakEnerg_FB_x | 9.421 | 0.002 | 12.071 | <0.001 | 0.979 | 0.418 | 0.028 |
| cov_a | 1.463 | 0.227 | 1.341 | 0.252 | 1.493 | 0.202 | 0.844 |
| Balance Task | | | | | | | |
| Features | F | *P* | F | *P* | F | *P* | n=189 |
| Power_MF_trem_z | 29.608 | <0.001 | 1.096 | 0.357 | 0.549 | 0.7 | 0.192 |
| RHL_trem_z | 19.372 | <0.001 | 1.848 | 0.117 | 1.712 | 0.145 | 0.349 |
| RHL_trem_a | 29.005 | <0.001 | 1.442 | 0.217 | 0.254 | 0.907 | 0.108 |
| Power_trem_z | 13.015 | <0.001 | 2.885 | 0.021 | 0.526 | 0.717 | 0.143 |
| Power_LF_trem_z | 15.785 | <0.001 | 5.59 | <0.001 | 0.901 | 0.462 | 0.614 |
| CFREQ_post_z | 9.483 | 0.002 | 1.532 | 0.19 | 0.772 | 0.544 | 0.066 |
| iqr_post_y | 33.914 | <0.001 | 10.249 | <0.001 | 0.609 | 0.656 | 0.245 |
| mean_post_y | 5.178 | 0.023 | 11.917 | <0.001 | 0.328 | 0.859 | <0.001 |
| min_post_y | 20.291 | <0.001 | 16.579 | <0.001 | 0.882 | 0.474 | 0.555 |
| F50_post_x | 0.975 | 0.324 | 0.631 | 0.641 | 1.372 | 0.241 | 0.285 |
| F50_post_a | 0.048 | 0.827 | 0.607 | 0.657 | 1.1 | 0.355 | 0.561 |
| rms_post_a | 16.826 | <0.001 | 5.588 | <0.001 | 0.869 | 0.482 | 0.603 |
| F50_post_x_z | 0.006 | 0.938 | 0.157 | 0.96 | 1.421 | 0.224 | 0.106 |
| TotalPower_post_z | 2.378 | 0.124 | 3.339 | 0.01 | 0.404 | 0.806 | 0.684 |
| TotalPower_post_y | 14.926 | <0.001 | 7.7 | <0.001 | 0.091 | 0.985 | 0.958 |
| Voice Task | | | | | | | |
| Features | F | *P* | F | *P* | F | *P* | n=280 |
| mean_gqc | 92.952 | <0.001 | 3.284 | 0.011 | 2.551 | 0.037 | 0.108 |
| c_mean_MFCC1 | 38.813 | <0.001 | 24.89 | <0.001 | 1.836 | 0.119 | 0.611 |
| p95_gqc | 59.181 | <0.001 | 1.531 | 0.19 | 1.706 | 0.146 | 0.016 |
| c_mean_MFCC10 | 22.916 | <0.001 | 0.796 | 0.528 | 0.586 | 0.673 | 0.257 |
| c_mean_MFCC12 | 10.811 | 0.001 | 2.232 | 0.063 | 1.304 | 0.266 | 0.012 |
| c_mean_MFCC7 | 13.614 | <0.001 | 1.009 | 0.401 | 1.73 | 0.14 | 0.182 |
| shdb | 7.787 | 0.005 | 1.985 | 0.094 | 0.538 | 0.708 | 0.375 |
| c_mean_MFCC5 | 7.142 | 0.008 | 2.813 | 0.024 | 0.198 | 0.939 | 0.084 |
| c_mean_MFCC9 | 1.264 | 0.261 | 1.554 | 0.184 | 0.205 | 0.936 | 0.186 |
| c_mean_MFCC8 | 1.373 | 0.242 | 1.375 | 0.24 | 0.552 | 0.698 | 0.676 |
| c_mean_MFCC6 | 0.936 | 0.334 | 2.035 | 0.087 | 1.011 | 0.4 | 0.982 |
| c_mean_MFCC3 | 1.886 | 0.17 | 7.87 | <0.001 | 6.754 | <0.001 | 0.249 |
| Tapping Task | | | | | | | |
|  | F | *P* | F | *P* | F | *P* | n=338 |
| numberTaps | 539.151 | <0.001 | 123.309 | <0.001 | 4.444 | 0.001 | <0.001 |
| buttonNoneFreq | 50.866 | <0.001 | 4.93 | <0.001 | 2.344 | 0.052 | 0.929 |
| mad_TapInter | 87.645 | <0.001 | 0.721 | 0.578 | 4.172 | 0.002 | 0.593 |
| median_DriftRight | 8.987 | 0.003 | 9.747 | <0.001 | 6.351 | <0.001 | 0.88 |
| mad_DriftRight | 8.9 | 0.003 | 10.605 | <0.001 | 6.353 | <0.001 | 0.846 |
| median_DriftLeft | 4.491 | 0.034 | 3.107 | 0.015 | 2.769 | 0.026 | 0.683 |
| min_TapInter | 44.616 | <0.001 | 11.437 | <0.001 | 0.388 | 0.818 | 0.226 |
| sd_DriftRight | 4.338 | 0.037 | 6.689 | <0.001 | 3.516 | 0.007 | 0.279 |
| iqr_TapInter | 53.591 | <0.001 | 1.054 | 0.378 | 2.553 | 0.037 | 0.319 |
| mad_DriftLeft | 2.594 | 0.108 | 11.145 | <0.001 | 6.06 | <0.001 | 0.548 |
| cv_TapInter | 8.054 | 0.005 | 6.815 | <0.001 | 1.702 | 0.146 | 0.232 |
| corXY | 5.935 | 0.015 | 21.815 | <0.001 | 2.541 | 0.038 | 0.874 |

HC- Healthy Controls, PD- Parkinson’s Disease, M- Male, F- Female, S.D- Standard Deviation, Gait Task: FreezeInd- Freeze Index, PeakEnerg- Peak of Energy, frec_peak- Frequency at the Peak of Energy, skew- Skewness, iqr- Interquartile Range, cov- Coefficient of Variation, zcr- Zero-Crossing Rate, kur-Kurtosis, LB- Locomotor Band, FB- Freezing Band, a- Accelerometer Average Signal, x- Accelerometer Mediolateral Signal, y- Accelerometer Vertical Signal, z- Accelerometer Anteroposterior Signal, Balance Task: PeakEnergy - Peak of energy, TotalPower- Energy between .15-3.5 Hz, Power- Energy between 3.5-15Hz, rms- Root Mean Square, F50- Frequency Containing 50% of Total Power, FRQD- Frequency of Dispersion of the Power Spectrum, iqr- Interquartile Range, min- Minimum Value, CFREQ- Centroidal Frequency, RHL- Ratio Between Power in High Frequency and Low Frequency, MF- Medium Frequency (4-7Hz), VHF- Very High Frequency (>7Hz) , HF- Hight Frequency (>4Hz), LF- Low Frequency (0.15-3.5Hz), trem- Tremor, post- Postural, a- Accelerometer Average Signal, x- Accelerometer Mediolateral Signal, y- Accelerometer Vertical Signal, z- Accelerometer Anteroposterior Signal, Hz- Hertz, Voice Task: c_mean_MFCC1–12- Mean Value of Mel Frequency Cepstral Coefficients 1-12, shbd- Shimmer, gqc- Glottis Quotient Close, p95- 95th Percentile, Tapping Task: Tapping Task: iqr- Interquartile Range, TapInter- Tap Interval, buttonNoneFreq: Frequency of Tapping Outside the Button, numberTaps- Number of Taps, DriftRight- Right Drift, corXY- Correlation of X and Y Positions, DriftLeft- Left Drift, mad- Median Absolute Deviation, min- Minimum, cv- Coefficient, Sd- Standard Deviation.

**Table S6.** Results from Mann-Whitney U test, Cohen’s d and Median ICC for Gait task. Median ICC across different time points and different repetitions for PD and HC.

| Feature Name | Mann-Whitney  U test  *P Value* | \|Cohens’d\| | Median ICC | | | |
| --- | --- | --- | --- | --- | --- | --- |
|  |  |  | Time Point | | Repetition | |
|  |  |  | HC | PD | HC | PD |
| frec_peak_LB_a | <0.001 | 0.3564 | 0.3213 | 0.2922 | 0.4335 | 0.3019 |
| FreezeInd_z | <0.001 | 0.281 | 0.3706 | 0.2629 | 0.3807 | 0.3129 |
| iqr_x | <0.001 | 0.2801 | 0.2242 | 0.2217 | 0.2521 | 0.2769 |
| MSI | <0.001 | 0.2619 | 0.15 | 0.2869 | 0.24 | 0.231 |
| numSteps | <0.001 | 0.2295 | 0.1417 | 0.2519 | 0.218 | 0.2265 |
| median_acc | <0.001 | 0.225 | 0.2452 | 0.1278 | 0.1907 | 0.1501 |
| PeakEnerg_LB_x | <0.001 | 0.2223 | 0.2338 | 0.2157 | 0.2458 | 0.2515 |
| min_z | 0.001 | 0.2205 | 0.2101 | 0.171 | 0.2324 | 0.2106 |
| ApEn_pos_z | 0.003 | 0.2127 | 0.0521 | 0.1015 | 0.0599 | 0.1263 |
| Power_LB_x | 0.001 | 0.2105 | 0.2141 | 0.2541 | 0.2726 | 0.3131 |
| Power_FB_z | <0.001 | 0.2088 | 0.4055 | 0.2066 | 0.3187 | 0.317 |
| ApEn_pos_a | 0.006 | 0.2047 | 0.0239 | 0.1807 | 0.0461 | 0.1396 |
| mean_a | 0.029 | 0.1903 | 0.138 | 0.18 | 0.1776 | 0.2488 |
| iqr_acc | 0.003 | 0.1833 | 0.0641 | 0.2097 | 0.1114 | 0.2566 |
| mean_acc | 0.004 | 0.1818 | -0.0834 | 0.0573 | 0.0267 | -0.0259 |
| rms_acc | 0.012 | 0.1736 | -0.0149 | 0.1103 | 0.1055 | 0.1264 |
| rms_y | 0.013 | 0.1707 | 0.1466 | 0.1984 | 0.2373 | 0.228 |
| PeakEnerg_LB_a | <0.001 | 0.1674 | 0.3301 | 0.3331 | 0.3356 | 0.3769 |
| RatioPower_y | 0.029 | 0.1646 | 0.1631 | 0.2649 | 0.2408 | 0.2981 |
| kur_pos_z | 0.003 | 0.1628 | 0.1048 | 0.055 | 0.0521 | 0.0607 |
| frec_peak_FB_vel_10 | 0.004 | 0.1609 | 0.0693 | 0.0929 | 0.0728 | 0.1018 |
| iqr_z | 0.005 | 0.1603 | 0.2659 | 0.2444 | 0.2908 | 0.3284 |
| skew_z | 0.003 | 0.1589 | 0.3317 | 0.1838 | 0.3627 | 0.2843 |
| frec_peak_FB_a | 0.001 | 0.1574 | 0.1287 | 0.1786 | 0.1707 | 0.1602 |
| median_a | 0.023 | 0.157 | 0.1741 | 0.262 | 0.2448 | 0.3192 |
| FreezeInd_x | 0.003 | 0.1568 | 0.2882 | 0.2689 | 0.3965 | 0.255 |
| Power_LB_z | <0.001 | 0.1553 | 0.2284 | 0.2034 | 0.3033 | 0.2472 |
| max_acc | 0.022 | 0.1538 | 0.1236 | 0.1021 | 0.1352 | 0.133 |
| zcr_x | 0.002 | 0.1503 | 0.3311 | 0.3121 | 0.4098 | 0.3866 |
| Power_LB_y | 0.036 | 0.1424 | 0.2764 | 0.3183 | 0.3445 | 0.3732 |
| Power_FB_acc | 0.019 | 0.1416 | 0.1886 | 0.1934 | 0.1689 | 0.2459 |
| kur_pos_a | 0.034 | 0.1407 | 0.0457 | 0.0972 | 0.0852 | 0.1431 |
| iqr_y | 0.019 | 0.1357 | 0.2813 | 0.3066 | 0.3125 | 0.367 |
| FreezeInd_a | 0.039 | 0.1321 | 0.2859 | 0.2453 | 0.321 | 0.2287 |
| Power_FB_vel | 0.009 | 0.1301 | 0.0782 | 0.132 | 0.1196 | 0.1779 |
| kur_x | 0.029 | 0.1295 | 0.268 | 0.2131 | 0.3992 | 0.3336 |
| frec_peak_LB_z | 0.001 | 0.129 | 0.2095 | 0.3273 | 0.3098 | 0.3143 |
| COEFCEPS20_pos_y | 0.008 | 0.1284 | 0.0739 | 0.0609 | 0.0456 | 0.0069 |
| COEFCEPS7_z | 0.004 | 0.1243 | -0.0383 | 0.057 | -0.0183 | 0.038 |
| PeakEnerg_FB_x | 0.039 | 0.122 | 0.3281 | 0.1444 | 0.3445 | 0.2341 |
| skew_vel | 0.013 | 0.1213 | 0.1296 | 0.1193 | 0.0878 | 0.1066 |
| cov_vel | 0.013 | 0.1188 | 0.101 | 0.1169 | 0.0748 | 0.114 |
| COEFCEPS1_pos_x | 0.017 | 0.1173 | -0.0056 | -0.0537 | 0.0362 | 0.0325 |
| iqr_pos_z | 0.021 | 0.1137 | 0.0342 | 0.0343 | 0.0799 | 0.0666 |
| cov_a | 0.009 | 0.1129 | 0.4435 | 0.4726 | 0.4793 | 0.5028 |
| FreezeInd_pos_a | 0.022 | 0.1108 | 0.0767 | 0.0169 | 0.1282 | 0.0308 |
| COEFCEPS9_z | 0.04 | 0.1107 | -0.007 | -0.0297 | -0.0125 | -0.0148 |
| PeakEnerg_LB_z | 0.004 | 0.1076 | 0.2642 | 0.2013 | 0.2253 | 0.2781 |
| COEFCEPS8_z | 0.045 | 0.107 | 0.0796 | 0.0194 | -0.003 | -0.0192 |
| COEFCEPS6_z | 0.05 | 0.1062 | -0.1046 | 0.0624 | -0.0412 | 0.0281 |
| frec_peak_LB_y | <0.001 | 0.1048 | 0.175 | 0.1178 | 0.2701 | 0.1949 |
| ApEn_vel | 0.015 | 0.1028 | 0.1951 | 0.1016 | 0.1224 | 0.069 |
| ApEn_pos_x | 0.015 | 0.0968 | -0.0401 | 0.0387 | -0.0158 | 0.0714 |
| median_y | 0.043 | 0.0926 | 0.1698 | 0.2399 | 0.2648 | 0.3101 |
| COEFCEPS10_z | 0.036 | 0.0922 | -0.0703 | -0.0739 | 0.0019 | -0.0175 |
| PeakEnerg_LB_pos_z | <0.001 | 0.0904 | 0.0393 | 0.0388 | 0.0533 | 0.0057 |
| COEFCEPS1_pos_a | 0.048 | 0.0882 | 0.0508 | 0.0852 | 0.0656 | 0.0644 |
| zcr_pos_z | 0.049 | 0.0814 | 0.0262 | 0.1287 | 0.0571 | 0.1752 |
| RatioPower_pos_z | 0.002 | 0.075 | -0.0051 | 0.0112 | 0.0302 | -0.0045 |
| RatioPower_pos_a | 0.038 | 0.0726 | -0.0101 | -0.0107 | 0.0188 | -0.0098 |
| Power_FB_pos_z | 0.014 | 0.0721 | -0.0136 | 0.008 | 0.0269 | -0.0055 |
| ApEn_pos_y | 0.037 | 0.0656 | 0.088 | 0.121 | 0.0823 | 0.099 |
| kur_vel | 0.039 | 0.0587 | 0.2061 | 0.0857 | 0.0866 | 0.0433 |
| cov_acc | 0.048 | 0.037 | -0.0013 | -0.0008 | -0.0096 | -0.003 |
| min_pos_x | 0.021 | 0.0359 | -0.0302 | 0.0342 | -0.0335 | 0.0243 |
| min_pos_y | 0.023 | 0.0027 | -0.0083 | 0.0136 | -0.023 | -0.0153 |

PD- Parkinson’s Disease, HC- Healthy Control, frec_peak- Frequency at the Peak of Energy, FreezeInd- Freeze Index, iqr-Interquartile Range, MSI- Mean Stride Interval, numSteps- Number of Steps, PeakEnerg- Peak of Energy, ApEn- Entropy, rms- Root Mean Square, RatioPower - Sum of the Power in the Freezing and Locomotor Band, skew- Skewness, min- Minimum Value, cov- Coefficient of Variation, zcr- Zero-Crossing Rate, kur-Kurtosis, COEFCEPS (1-20)- Mel Frequency Cepstral Coefficients, ar- Coefficient of a 1st Order Autoregressive Model, LB- Locomotor Band, FB- Freezing Band, vel- Velocity, acc- Acceleration Along Path, a- Accelerometer Average Signal, x- Accelerometer Mediolateral Signal, y- Accelerometer Vertical Signal, z- Accelerometer Anteroposterior Signal.

**Table S7** Results from Mann-Whitney U test, Cohen’s d and Median ICC for features from Balance task. Median ICC across different time points and different repetitions for PD and HC. Features are selected based on Mann-Whitney U test that significantly differ between PD and HC at the first administration (baseline) (*P*<.05).

| Feature Name | *Mann-Whitney*  *U Test*  *P Value* | \|Cohens'd\| | Median ICC | | | |
| --- | --- | --- | --- | --- | --- | --- |
|  |  |  | Time Point | | Repetition | |
|  |  |  | HC | PD | HC | PD |
| Power_MF_trem_z | <0.001 | 0.337 | 0.2808 | 0.2956 | 0.3016 | 0.2844 |
| PeakEnerg_VHF_trem_x | 0.028 | 0.3129 | 0.2521 | 0.3188 | 0.159 | 0.2677 |
| PeakEnerg_VHF_trem_z | <0.001 | 0.2942 | 0.3225 | 0.2267 | 0.2318 | 0.205 |
| RHL_trem_z | <0.001 | 0.2679 | 0.2891 | 0.5605 | 0.2936 | 0.1331 |
| RHL_trem_a | 0.004 | 0.2517 | 0.4071 | 0.3245 | 0.4463 | 0.294 |
| Power_trem_y | <0.001 | 0.2441 | 0.0492 | 0.1998 | 0.1737 | 0.1535 |
| F95_post_y | <0.001 | 0.2402 | 0.1742 | 0.1308 | 0.2351 | 0.1838 |
| Power_trem_z | 0.006 | 0.2317 | 0.2601 | 0.2376 | 0.2824 | 0.2763 |
| median_trem_a | <0.001 | 0.2203 | 0.0973 | 0.1609 | 0.1475 | 0.1638 |
| Power_LF_trem_z | <0.001 | 0.2186 | 0.2016 | 0.1727 | 0.2414 | 0.1817 |
| CFREQ_post_z | 0.008 | 0.2057 | 0.3047 | 0.2654 | 0.3369 | 0.2716 |
| F95_post_x | 0.002 | 0.1956 | 0.1938 | 0.1679 | 0.2122 | 0.1707 |
| MFREQ_dist_x | <0.001 | 0.1915 | 0.0429 | 0.0269 | 0.0494 | 0.0648 |
| iqr_post_y | <0.001 | 0.1873 | 0.1721 | 0.2481 | 0.1837 | 0.2804 |
| mean_trem_a | <0.001 | 0.1851 | 0.1015 | 0.1805 | 0.1317 | 0.2164 |
| kur_trem_x | <0.001 | 0.1799 | 0.0749 | 0.0185 | 0.0634 | 0.0222 |
| F95_post_a | 0.012 | 0.1776 | 0.1597 | 0.1905 | 0.2026 | 0.2202 |
| ApEn_trem_x | <0.001 | 0.1757 | 0.0841 | 0.091 | 0.1253 | 0.0905 |
| zcr_post_y | <0.001 | 0.1733 | 0.2208 | 0.2434 | 0.2412 | 0.1457 |
| median_post_a | <0.001 | 0.1724 | 0.1195 | 0.0992 | 0.1907 | 0.1524 |
| iqr_trem_x | 0.002 | 0.1699 | 0.16 | 0.1447 | 0.1766 | 0.1522 |
| FD_CC_dist_x_z | <0.001 | 0.1638 | 0.0852 | -0.0044 | 0.0214 | 0.0247 |
| F50_post_y | 0.001 | 0.1631 | 0.1875 | 0.209 | 0.2339 | 0.2354 |
| Power_LF_trem_x | 0.01 | 0.1608 | 0.1362 | 0.1831 | 0.2339 | 0.1794 |
| range_trem_y | <0.001 | 0.1585 | 0.1733 | 0.0855 | 0.2059 | 0.1177 |
| MVELO_dist_x | 0.006 | 0.1573 | 0.1408 | 0.1285 | 0.2058 | 0.1832 |
| mean_post_y | 0.007 | 0.1532 | 0.3668 | 0.2441 | 0.3952 | 0.2764 |
| min_post_y | <0.001 | 0.1521 | 0.2216 | 0.2646 | 0.2126 | 0.2841 |
| iqr_post_x | 0.007 | 0.1449 | 0.1677 | 0.1459 | 0.1918 | 0.1951 |
| kur_post_x | 0.002 | 0.1391 | 0.0707 | 0.0247 | 0.0703 | 0.0539 |
| rms_trem_a | 0.004 | 0.1372 | 0.1666 | 0.152 | 0.1892 | 0.2266 |
| ApEn_post_a | 0.004 | 0.1361 | 0.0435 | 0.1382 | 0.1055 | 0.1037 |
| skew_post_a | <0.001 | 0.1317 | 0.0291 | 0.1097 | 0.0483 | 0.0813 |
| AREA_CC_dist_x_z | 0.013 | 0.1306 | 0.1307 | 0.0011 | 0.0632 | 0.0322 |
| FD_dist_x_z | 0.003 | 0.1303 | 0.0781 | -0.0472 | 0.019 | 0.0282 |
| cov_trem_a | <0.001 | 0.1289 | 0.0988 | 0.032 | 0.1202 | 0.0325 |
| TotalPower_post_x_z | 0.011 | 0.1277 | 0.156 | 0.0493 | 0.1804 | 0.0726 |
| MFREQ_dist_x_z | 0.001 | 0.1261 | 0.0836 | -0.0104 | 0.0415 | 0.0331 |
| max_post_y | <0.001 | 0.1245 | 0.1192 | 0.2187 | 0.2363 | 0.2089 |
| F50_post_x | <0.001 | 0.1243 | 0.2149 | 0.253 | 0.1823 | 0.29 |
| cov_post_a | 0.009 | 0.1199 | 0.0641 | 0.0677 | 0.0992 | 0.0634 |
| range_trem_x | 0.05 | 0.1195 | 0.1877 | 0.2015 | 0.2212 | 0.1329 |
| ApEn_trem_y | 0.001 | 0.1146 | 0.0886 | 0.0736 | 0.1512 | 0.1158 |
| FD_CE_dist_x_z | 0.007 | 0.1055 | 0.1519 | 0.0453 | 0.0895 | 0.0774 |
| mean_post_z | 0.031 | 0.0994 | 0.1935 | 0.1906 | 0.1498 | 0.1938 |
| F50_post_a | 0.003 | 0.0988 | 0.2095 | 0.2191 | 0.243 | 0.2912 |
| Power_LF_trem_a | 0.005 | 0.0982 | 0.217 | 0.0347 | 0.185 | 0.0634 |
| max_post_z | 0.027 | 0.0939 | 0.207 | 0.1773 | 0.1862 | 0.2413 |
| rms_post_a | 0.028 | 0.0925 | 0.2293 | 0.1188 | 0.2554 | 0.1798 |
| F50_post_x_z | 0.006 | 0.0882 | 0.2172 | 0.2453 | 0.2351 | 0.3015 |
| FREQD_post_x | 0.005 | 0.0848 | 0.1709 | 0.2444 | 0.1828 | 0.2444 |
| TotalPower_post_z | 0.014 | 0.0837 | 0.226 | 0.1239 | 0.2691 | 0.192 |
| FREQD_post_x_z | 0.019 | 0.0811 | 0.1243 | 0.1909 | 0.1926 | 0.1707 |
| TotalPower_post_y | <0.001 | 0.0805 | 0.132 | 0.1737 | 0.216 | 0.2851 |
| kur_post_a | <0.001 | 0.0717 | 0.0227 | 0.0732 | 0.0094 | 0.0578 |
| max_trem_z | 0.038 | 0.0709 | 0.0628 | 0.0578 | 0.0982 | 0.1206 |
| jerk_post_y | <0.001 | 0.0421 | 0.0474 | 0.0484 | 0.181 | 0.1174 |
| kur_trem_a | 0.012 | 0.0349 | 0.0081 | 0.0034 | 0.0014 | -0.0098 |
| kur_trem_y | 0.022 | 0.0233 | 0.0305 | -0.0257 | 0.0612 | -0.0137 |

PD- Parkinson’s Disease, HC- Healthy Control, PeakEnergy - Peak of energy, TotalPower- Energy between 15-3.5 Hz, rms- Root Mean Square, F50- Frequency Containing 50% of Total Power, F95- Frequency containing 95% of the total power, FRQD- Frequency of Dispersion of the Power Spectrum, MFREQ- Mean Frequency, iqr- Interquartile Range, kur- Kurtosis, zcr- Zero-Crossing Rate, ApEn- Entropy, skew- Skewness, jerk- Average jerk, MVELO- Mean velocity, FD- Fractal Dimension, FD_CE- Fractal Dimension based on the 95% Confidence Ellipse Area, min- Minimum Value, CFREQ- Centroidal Frequency, RHL- Ratio Between Power in High Frequency and Low Frequency, dist- Distance, MF- Medium Frequency (4-7Hz), VHF- Very High Frequency (>7Hz), HF- Hight Frequency (>4Hz), LF- Low Frequency (0.15-3.5Hz), trem- Tremor, post- Postural, a- Accelerometer Average Signal, x- Accelerometer Mediolateral Signal, y- Accelerometer Vertical Signal, z- Accelerometer Anteroposterior Signal, Hz- Hertz.

**Table s8.** Results from Mann-Whitney U test, Cohen’s d and Median ICC for features from Voice task. Median ICC across different time points and different repetitions for PD and HC. Features are selected based on Mann-Whitney U test that significantly differ between PD and HC at the first administration (baseline) (*P*<.05).

| Feature Name | Mann-Whitney  U Test  *P Value* | \|Cohens'd\| | Median ICC | | | |
| --- | --- | --- | --- | --- | --- | --- |
|  |  |  | Time Point | | Repetition | |
|  |  |  | HC | PD | HC | PD |
| mean_gqc | <0.001 | 0.5049 | 0.6737 | 0.7093 | 0.6797 | 0.7476 |
| p5_gqc | <0.001 | 0.412 | 0.3308 | 0.37 | 0.3169 | 0.4305 |
| c_mean_MFCC1 | <0.001 | 0.4028 | 0.5158 | 0.4304 | 0.5874 | 0.5071 |
| p95_gqc | <0.001 | 0.354 | 0.6699 | 0.6128 | 0.6779 | 0.6494 |
| std_tkeo | <0.001 | 0.3033 | 0.4022 | 0.3972 | 0.4577 | 0.4393 |
| p95_tkeo | <0.001 | 0.2919 | 0.318 | 0.3387 | 0.3596 | 0.397 |
| c_std_d11 | <0.001 | 0.278 | 0.3747 | 0.3887 | 0.4209 | 0.4159 |
| hnr_std | <0.001 | 0.2738 | 0.2193 | 0.2973 | 0.2553 | 0.3413 |
| c_std_d12 | <0.001 | 0.2717 | 0.3335 | 0.4011 | 0.4497 | 0.4253 |
| p75_tkeo | <0.001 | 0.2679 | 0.4344 | 0.2648 | 0.4264 | 0.3476 |
| c_std_d13 | <0.001 | 0.2634 | 0.3766 | 0.3868 | 0.4272 | 0.4303 |
| c_std_d8 | <0.001 | 0.2491 | 0.3019 | 0.345 | 0.3748 | 0.3711 |
| c_std_d9 | <0.001 | 0.2445 | 0.3108 | 0.3664 | 0.3589 | 0.3817 |
| c_mean_MFCC10 | <0.001 | 0.2403 | 0.5648 | 0.6197 | 0.6255 | 0.6454 |
| c_std_d10 | <0.001 | 0.2401 | 0.3423 | 0.389 | 0.4058 | 0.4132 |
| c_std_d7 | <0.001 | 0.2309 | 0.3589 | 0.3308 | 0.3779 | 0.3758 |
| c_std_dd11 | <0.001 | 0.2282 | 0.4119 | 0.4281 | 0.4627 | 0.4498 |
| c_std_d5 | <0.001 | 0.2196 | 0.3335 | 0.2868 | 0.3318 | 0.3435 |
| c_std_d14 | <0.001 | 0.2193 | 0.3474 | 0.3912 | 0.3987 | 0.4291 |
| c_std_d6 | <0.001 | 0.2186 | 0.3699 | 0.3492 | 0.3717 | 0.3718 |
| c_mean_MFCC12 | <0.001 | 0.2154 | 0.4902 | 0.433 | 0.5362 | 0.4977 |
| c_std_dd5 | <0.001 | 0.2048 | 0.373 | 0.369 | 0.4207 | 0.4006 |
| c_std_d3 | <0.001 | 0.202 | 0.2891 | 0.3135 | 0.2862 | 0.3391 |
| c_std_d4 | <0.001 | 0.2008 | 0.3006 | 0.3027 | 0.3129 | 0.3435 |
| c_std_dd10 | <0.001 | 0.1976 | 0.4096 | 0.4436 | 0.4759 | 0.4647 |
| c_std_dd9 | <0.001 | 0.1942 | 0.3812 | 0.4232 | 0.4336 | 0.4327 |
| c_std_dd8 | <0.001 | 0.1932 | 0.3741 | 0.4008 | 0.454 | 0.4177 |
| c_std_dd12 | <0.001 | 0.1923 | 0.4006 | 0.446 | 0.4961 | 0.4601 |
| c_std_MFCC1 | 0.026 | 0.1883 | 0.2528 | 0.251 | 0.2488 | 0.3304 |
| c_std_MFCC5 | 0.001 | 0.1853 | 0.2922 | 0.2074 | 0.3001 | 0.2461 |
| c_std_dd7 | <0.001 | 0.1842 | 0.3847 | 0.3989 | 0.4603 | 0.4339 |
| c_std_dd13 | <0.001 | 0.1821 | 0.3979 | 0.4326 | 0.4806 | 0.4675 |
| c_std_dd6 | <0.001 | 0.182 | 0.3975 | 0.3964 | 0.4414 | 0.4133 |
| DFA | <0.001 | 0.1792 | 0.0577 | 0.0833 | 0.0365 | 0.1416 |
| c_mean_0th | 0.002 | 0.1675 | 0.1974 | 0.2609 | 0.2692 | 0.2913 |
| c_mean_d13 | <0.001 | 0.1673 | 0.1084 | 0.1388 | 0.1036 | 0.127 |
| fm | 0.001 | 0.1648 | 0.3646 | 0.3997 | 0.3755 | 0.3536 |
| c_std_dd4 | 0.002 | 0.1631 | 0.3667 | 0.3165 | 0.3905 | 0.3851 |
| c_mean_MFCC7 | 0.001 | 0.1615 | 0.5329 | 0.5577 | 0.6043 | 0.5592 |
| c_mean_d4 | 0.002 | 0.1597 | 0.0228 | 0.1646 | 0.0771 | 0.1991 |
| shdb | 0.03 | 0.1552 | 0.3822 | 0.4779 | 0.5244 | 0.523 |
| c_std_dd3 | 0.001 | 0.1495 | 0.3525 | 0.3232 | 0.3634 | 0.3412 |
| ApEn_f0 | <0.001 | 0.148 | 0.0964 | 0.145 | 0.1605 | 0.2034 |
| c_std_dd14 | 0.006 | 0.1447 | 0.3701 | 0.4477 | 0.4625 | 0.4762 |
| c_mean_d3 | 0.001 | 0.1344 | 0.1441 | 0.1025 | 0.1508 | 0.1771 |
| c_std_MFCC11 | 0.016 | 0.1323 | 0.231 | 0.2937 | 0.3196 | 0.3401 |
| c_std_d1 | 0.031 | 0.1292 | 0.2985 | 0.3704 | 0.2786 | 0.3606 |
| c_std_MFCC6 | 0.005 | 0.1261 | 0.2722 | 0.2996 | 0.3064 | 0.333 |
| c_mean_MFCC5 | 0.008 | 0.1228 | 0.5375 | 0.5294 | 0.5745 | 0.5899 |
| c_mean_MFCC9 | 0.004 | 0.1209 | 0.4986 | 0.4885 | 0.5747 | 0.5466 |
| c_std_MFCC9 | 0.014 | 0.1172 | 0.2365 | 0.2543 | 0.285 | 0.2889 |
| rpde | 0.002 | 0.1161 | 0.3277 | 0.3635 | 0.3636 | 0.3956 |
| c_std_MFCC7 | 0.041 | 0.1087 | 0.2765 | 0.3007 | 0.3085 | 0.3234 |
| c_mean_MFCC8 | 0.048 | 0.0999 | 0.4376 | 0.5004 | 0.5097 | 0.524 |
| c_std_MFCC8 | 0.028 | 0.0934 | 0.1816 | 0.3283 | 0.2736 | 0.3469 |
| c_mean_MFCC6 | 0.019 | 0.081 | 0.5523 | 0.5853 | 0.587 | 0.5872 |
| c_mean_d2 | 0.002 | 0.0781 | 0.1124 | 0.1832 | 0.1251 | 0.2258 |
| c_mean_d6 | 0.039 | 0.0725 | 0.1365 | 0.1728 | 0.1582 | 0.1926 |
| c_mean_MFCC3 | 0.02 | 0.0706 | 0.4717 | 0.5061 | 0.5486 | 0.5317 |
| c_mean_d1 | 0.005 | 0.0705 | 0.1218 | 0.1949 | 0.1002 | 0.2329 |

PD- Parkinson’s Disease, HC- Healthy Control, c_mean- Mean of the MFCCs Coefficients, log-Energy of the Signal and the First and Second Derivatives of the MFCCs, MFCC- Mel Frequency Cepstral Coefficients, c_std- Standard Deviation of the MFCCs Coefficients, gqc- Glottis Quotient Close, fm- Frequency Modulation, std - Standard Deviation, tkeo- Teager Kaiser Energy Operator, p5- 5th percentile, p75- 75th Percentile, p95- 95th Percentile, shbd- Shimmer, hnr- Harmonic to Noise Ratio, d- Delta, d-d- Delta-Delta, DFA- Detrended Fluctuation Analysis, f0- Fundamental Frequency, ApEn- Pitch Period Entropy, rpde- Recurrence Period Density Entropy, T- Period.

**Table S9.** Results from Mann-Whitney U test, Cohen’s d and Median ICC for features from Tapping task. Median ICC across different time points and different repetitions for PD and HC. Features are selected based on Mann-Whitney U test that significantly differ between PD and HC at the first administration (baseline) (*P*<.05).

| Feature Name | Mann-Whitney  U test  *P Value* | \|Cohens'd \| | Median ICC | | | |
| --- | --- | --- | --- | --- | --- | --- |
|  |  |  | Time Point | | Repetition | |
|  |  |  | HC | PD | HC | PD |
| numberTaps | <0.001 | 1.179 | 0.6896 | 0.6411 | 0.6895 | 0.7004 |
| max_TapInter | <0.001 | 0.6177 | 0.1792 | 0.2758 | 0.1838 | 0.2909 |
| range_TapInter | <0.001 | 0.5821 | 0.1297 | 0.2741 | 0.1628 | 0.2809 |
| ar2_TapInter | <0.001 | 0.5322 | 0.3102 | 0.2777 | 0.3286 | 0.3243 |
| ar1_TapInter | <0.001 | 0.5282 | 0.3039 | 0.2938 | 0.3193 | 0.3217 |
| sd_TapInter | <0.001 | 0.5154 | 0.2779 | 0.2258 | 0.3023 | 0.3459 |
| buttonNoneFreq | <0.001 | 0.4455 | 0.3076 | 0.3658 | 0.3453 | 0.4613 |
| mad_TapInter | <0.001 | 0.3701 | 0.3294 | 0.305 | 0.3488 | 0.3751 |
| median_DriftRight | <0.001 | 0.3542 | 0.6796 | 0.5393 | 0.4578 | 0.5345 |
| mad_DriftRight | <0.001 | 0.3468 | 0.4492 | 0.4259 | 0.466 | 0.4822 |
| median_DriftLeft | <0.001 | 0.2912 | 0.51 | 0.5269 | 0.5202 | 0.5292 |
| min_TapInter | <0.001 | 0.2857 | 0.2751 | 0.3967 | 0.4364 | 0.4605 |
| sd_DriftRight | <0.001 | 0.2842 | 0.4088 | 0.2833 | 0.4066 | 0.3227 |
| iqr_TapInter | <0.001 | 0.2785 | 0.4353 | 0.3046 | 0.4745 | 0.3506 |
| mad_DriftLeft | <0.001 | 0.2725 | 0.4603 | 0.4723 | 0.4801 | 0.4591 |
| sd_DriftLeft | <0.001 | 0.2694 | 0.3654 | 0.4028 | 0.3932 | 0.2981 |
| skew_TapInter | <0.001 | 0.2315 | 0.1461 | 0.1855 | 0.1614 | 0.2029 |
| skew_DriftLeft | <0.001 | 0.208 | 0.0637 | 0.1046 | 0.092 | 0.1149 |
| kur_DriftLeft | <0.001 | 0.1913 | 0.0299 | 0.063 | 0.0924 | 0.0827 |
| kur_DriftRight | <0.001 | 0.1842 | 0.1257 | 0.0258 | 0.1567 | 0.0796 |
| kur_TapInter | <0.001 | 0.1695 | 0.092 | 0.1072 | 0.1043 | 0.1117 |
| cv_TapInter | <0.001 | 0.1586 | 0.4259 | 0.4212 | 0.4716 | 0.4547 |
| corXY | 0.001 | 0.114 | 0.624 | 0.5737 | 0.5804 | 0.5324 |
| tkeo_TapInter | <0.001 | 0.0905 | 0.0437 | 0.0275 | 0.0439 | 0.0988 |
| cv_DriftLeft | 0.014 | 0.0113 | 0.0981 | 0.135 | 0.1078 | 0.162 |

PD- Parkinson’s Disease, HC- Healthy Control, iqr- Interquartile Range, TapInter- Tap Interval, buttonNoneFreq: Frequency of Tapping Outside the Button, numberTaps- Number of Taps, DriftRight- Right Drift, corXY- Correlation of X and Y Positions, DriftLeft- Left Drift, mad- Median Absolute Deviation, min- Minimum, max- Maximum, skew- Skewness, kur- Kurtosis, teko- Teager-Kaiser Energy Operator, cv- Coefficient, Sd- Standard Deviation, ar (1-2)- Coefficient of an Autoregressive Model at Lag (1-2).

**Table S10.** Results from an analysis of ANOVA for repeated measurements on the features from Gait task. Features are selected based on Mann-Whitney U test that significantly differ between PD and HC at the first administration (baseline) (*P*<.05).

| Feature Name | Medication | | diagnosis  (PD, HC) | | Repetition | | Diagnosis × Repetition | |
| --- | --- | --- | --- | --- | --- | --- | --- | --- |
|  | F | *P* | F | *P* | F | *P* | F | *P* |
| frec_peak_LB_a | 1.03 | 0.358 | 23.202 | <0.001 | 0.767 | 0.547 | 0.276 | 0.894 |
| FreezeInd_z | 0.864 | 0.422 | 3.877 | 0.049 | 0.665 | 0.616 | 1.012 | 0.4 |
| iqr_x | 0.348 | 0.706 | 1.502 | 0.221 | 12.892 | <0.001 | 1.77 | 0.132 |
| MSI | 0.637 | 0.529 | 14.831 | <0.001 | 0.61 | 0.655 | 0.113 | 0.978 |
| numSteps | 0.696 | 0.499 | 17.274 | <0.001 | 1.038 | 0.386 | 0.067 | 0.992 |
| median_acc | 0.074 | 0.928 | 9.673 | 0.002 | 2.833 | 0.023 | 1.262 | 0.283 |
| PeakEnerg_LB_x | 0.104 | 0.901 | 0.003 | 0.954 | 12.381 | <0.001 | 1.703 | 0.147 |
| min_z | 0.564 | 0.57 | 5.477 | 0.02 | 2.724 | 0.028 | 3.282 | 0.011 |
| ApEn_pos_z | 0.249 | 0.779 | 13.714 | <0.001 | 0.701 | 0.591 | 1.472 | 0.208 |
| Power_LB_x | 1.85 | 0.159 | 0.215 | 0.643 | 8.244 | <0.001 | 1.845 | 0.118 |
| Power_FB_z | 2.549 | 0.08 | 0.144 | 0.704 | 10.235 | <0.001 | 4.719 | <0.001 |
| ApEn_pos_a | 1.239 | 0.291 | 2.417 | 0.121 | 2.913 | 0.02 | 1.005 | 0.404 |
| mean_a | 3.781 | 0.024 | 1.053 | 0.305 | 14.267 | <0.001 | 4.027 | 0.003 |
| iqr_acc | 3.521 | 0.031 | 0.014 | 0.904 | 10.881 | <0.001 | 2.116 | 0.076 |
| mean_acc | 0.411 | 0.663 | 9.221 | 0.002 | 1.353 | 0.248 | 0.227 | 0.923 |
| rms_acc | 3.682 | 0.026 | 0.088 | 0.767 | 8.63 | <0.001 | 4.71 | <0.001 |
| rms_y | 3.048 | 0.049 | 0.433 | 0.511 | 6.438 | <0.001 | 4.419 | 0.001 |
| PeakEnerg_LB_a | 0.982 | 0.376 | 10.268 | 0.001 | 9.974 | <0.001 | 3.02 | 0.017 |
| RatioPower_y | 2.183 | 0.114 | 1.898 | 0.169 | 8.466 | <0.001 | 3.447 | 0.008 |
| kur_pos_z | 0.724 | 0.486 | 10.965 | <0.001 | 0.923 | 0.45 | 2.679 | 0.03 |
| frec_peak_FB_vel_10 | 0.34 | 0.712 | 7.353 | 0.007 | 3.544 | 0.007 | 1.017 | 0.397 |
| iqr_z | 0.215 | 0.806 | 0.109 | 0.742 | 6.849 | <0.001 | 1.102 | 0.354 |
| skew_z | 1.658 | 0.192 | 9.226 | 0.002 | 2.749 | 0.027 | 0.267 | 0.899 |
| frec_peak_FB_a | 1.067 | 0.345 | 10.735 | 0.001 | 1.114 | 0.348 | 0.357 | 0.839 |
| median_a | 2.875 | 0.058 | 1.003 | 0.317 | 13.362 | <0.001 | 2.814 | 0.024 |
| FreezeInd_x | 1.899 | 0.151 | 2.681 | 0.102 | 0.954 | 0.432 | 0.406 | 0.804 |
| Power_LB_z | 0.264 | 0.768 | 9.674 | 0.002 | 11.043 | <0.001 | 1.32 | 0.26 |
| max_acc | 0.167 | 0.846 | 2.783 | 0.096 | 1.306 | 0.265 | 3.298 | 0.011 |
| zcr_x | 0.546 | 0.58 | 4.987 | 0.026 | 4.387 | 0.002 | 0.827 | 0.508 |
| Power_LB_y | 1.865 | 0.156 | 0.256 | 0.613 | 2.985 | 0.018 | 0.792 | 0.53 |
| Power_FB_acc | 0.771 | 0.463 | 1.372 | 0.242 | 9.907 | <0.001 | 5.476 | <0.001 |
| kur_pos_a | 0.934 | 0.394 | 0.885 | 0.347 | 4.982 | <0.001 | 2.408 | 0.047 |
| iqr_y | 4.641 | 0.01 | 0 | 1.0 | 2.28 | 0.059 | 1.223 | 0.299 |
| FreezeInd_a | 0.105 | 0.9 | 2.124 | 0.146 | 1.102 | 0.354 | 0.584 | 0.674 |
| Power_FB_vel | 0.797 | 0.452 | 0.585 | 0.445 | 1.37 | 0.242 | 1.374 | 0.24 |
| kur_x | 0.984 | 0.375 | 2.438 | 0.119 | 0.418 | 0.796 | 1.969 | 0.097 |
| frec_peak_LB_z | 0.802 | 0.449 | 1.013 | 0.315 | 2.198 | 0.067 | 0.705 | 0.589 |
| COEFCEPS20_pos_y | 0.227 | 0.797 | 0 | 1.0 | 3.391 | 0.009 | 0.51 | 0.729 |
| COEFCEPS7_z | 0.11 | 0.896 | 1.377 | 0.241 | 0.548 | 0.7 | 2.725 | 0.028 |
| PeakEnerg_FB_x | 3.604 | 0.028 | 9.421 | 0.002 | 12.071 | <0.001 | 0.979 | 0.418 |
| skew_vel | 0.582 | 0.559 | 20.913 | <0.001 | 3.57 | 0.007 | 2.99 | 0.018 |
| cov_vel | 0.67 | 0.512 | 12.713 | <0.001 | 2.013 | 0.09 | 2.824 | 0.024 |
| COEFCEPS1_pos_x | 0.171 | 0.843 | 13.528 | <0.001 | 3.217 | 0.012 | 0.333 | 0.856 |
| iqr_pos_z | 0.65 | 0.523 | 3.476 | 0.063 | 6.916 | <0.001 | 0.898 | 0.464 |
| cov_a | 0.17 | 0.844 | 1.463 | 0.227 | 1.341 | 0.252 | 1.493 | 0.202 |
| FreezeInd_pos_a | 0.355 | 0.702 | 0 | 1.0 | 2.16 | 0.071 | 1.584 | 0.176 |
| COEFCEPS9_z | 0.153 | 0.858 | 0.365 | 0.546 | 0.749 | 0.559 | 1.771 | 0.132 |
| PeakEnerg_LB_z | 0.13 | 0.878 | 6.525 | 0.011 | 11.047 | <0.001 | 0.405 | 0.805 |
| COEFCEPS8_z | 0.086 | 0.918 | 0 | 1.0 | 0.996 | 0.408 | 2.3 | 0.057 |
| COEFCEPS6_z | 0.675 | 0.51 | 0.54 | 0.463 | 0.463 | 0.763 | 1.135 | 0.338 |
| frec_peak_LB_y | 0.396 | 0.674 | 1.887 | 0.17 | 3.161 | 0.013 | 0.116 | 0.977 |
| ApEn_vel | 0.54 | 0.583 | 18.003 | <0.001 | 2.215 | 0.065 | 2.301 | 0.056 |
| ApEn_pos_x | 0.846 | 0.43 | 11.025 | <0.001 | 1.571 | 0.179 | 2.071 | 0.082 |
| median_y | 0.556 | 0.574 | 0.268 | 0.605 | 2.994 | 0.018 | 0.024 | 0.999 |
| COEFCEPS10_z | 0.231 | 0.794 | 0.108 | 0.742 | 0.859 | 0.488 | 1.824 | 0.122 |
| PeakEnerg_LB_pos_z | 0.381 | 0.683 | 4.483 | 0.035 | 3.138 | 0.014 | 0.575 | 0.681 |
| COEFCEPS1_pos_a | 0.496 | 0.609 | 3.953 | 0.047 | 1.476 | 0.207 | 0.818 | 0.514 |
| zcr_pos_z | 0.546 | 0.58 | 0 | 1.0 | 0 | 1.0 | 5.182 | <0.001 |
| RatioPower_pos_z | 0.483 | 0.617 | 2.316 | 0.129 | 3.968 | 0.003 | 0.359 | 0.838 |
| RatioPower_pos_a | 1.356 | 0.259 | 1.389 | 0.239 | 7.854 | <0.001 | 0.326 | 0.861 |
| Power_FB_pos_z | 0.491 | 0.613 | 1.871 | 0.172 | 4.19 | 0.002 | 0.321 | 0.864 |
| ApEn_pos_y | 0.214 | 0.807 | 4.055 | 0.045 | 2.091 | 0.079 | 0.558 | 0.693 |
| kur_vel | 0.449 | 0.639 | 14.725 | <0.001 | 1.367 | 0.243 | 2.074 | 0.082 |
| cov_acc | 0.433 | 0.649 | 0.175 | 0.676 | 1.029 | 0.391 | 0.864 | 0.485 |
| min_pos_x | 4.193 | 0.016 | 0.692 | 0.406 | 2.735 | 0.027 | 0.908 | 0.458 |
| min_pos_y | 2.354 | 0.096 | 0.069 | 0.793 | 6.382 | <0.001 | 1.057 | 0.377 |

PD- Parkinson’s Disease, HC- Healthy Control, frec_peak- Frequency at the Peak of Energy, FreezeInd- Freeze Index, iqr-Interquartile Range, MSI- Mean Stride Interval, numSteps- Number of Steps, PeakEnerg- Peak of Energy, ApEn- Entropy, rms- Root Mean Square, RatioPower - Sum of the Power in the Freezing and Locomotor Band, skew- Skewness, min- Minimum Value, cov- Coefficient of Variation, zcr- Zero-Crossing Rate, kur-Kurtosis, COEFCEPS (1-20)- Mel Frequency Cepstral Coefficients, ar- Coefficient of a 1st Order Autoregressive Model, LB- Locomotor Band, FB- Freezing Band, vel- Velocity, acc- Acceleration Along Path, a- Accelerometer Average Signal, x- Accelerometer Mediolateral Signal, y- Accelerometer Vertical Signal, z- Accelerometer Anteroposterior Signal.

**Table S11.** Results from an analysis of ANOVA for repeated measurements on the features from Balance task. Features are selected based on Mann-Whitney U test that significantly differ between PD and HC at the first administration (baseline) (*P*<.05).

| Feature Name | Medication | | Diagnosis  (PD, HC) | | Repetition | | diagnosis × Repetition | |
| --- | --- | --- | --- | --- | --- | --- | --- | --- |
|  | F | *P* | F | *P* | F | *P* | F | *P* |
| Power_MF_trem_z | 1.657 | 0.192 | 29.608 | <0.001 | 1.096 | 0.357 | 0.549 | 0.7 |
| PeakEnerg_VHF_trem_x | 0.643 | 0.526 | 31.861 | <0.001 | 0.165 | 0.956 | 0.345 | 0.848 |
| PeakEnerg_VHF_trem_z | 0.934 | 0.394 | 26.313 | <0.001 | 0.927 | 0.447 | 0.775 | 0.542 |
| RHL_trem_z | 1.055 | 0.349 | 19.372 | <0.001 | 1.848 | 0.117 | 1.712 | 0.145 |
| RHL_trem_a | 2.235 | 0.108 | 29.005 | <0.001 | 1.442 | 0.217 | 0.254 | 0.907 |
| Power_trem_y | 0.618 | 0.54 | 25.883 | <0.001 | 5.182 | <0.001 | 0.295 | 0.882 |
| F95_post_y | 0.337 | 0.714 | 17.64 | <0.001 | 6.648 | <0.001 | 0.241 | 0.915 |
| Power_trem_z | 1.956 | 0.143 | 13.015 | <0.001 | 2.885 | 0.021 | 0.526 | 0.717 |
| median_trem_a | 1.993 | 0.138 | 41.154 | <0.001 | 0.738 | 0.566 | 0.471 | 0.757 |
| Power_LF_trem_z | 0.488 | 0.614 | 15.785 | <0.001 | 5.59 | <0.001 | 0.901 | 0.462 |
| CFREQ_post_z | 2.744 | 0.066 | 9.483 | 0.002 | 1.532 | 0.19 | 0.772 | 0.544 |
| F95_post_x | 1.179 | 0.309 | 3.449 | 0.064 | 3.528 | 0.007 | 0.317 | 0.867 |
| MFREQ_dist_x | 0.153 | 0.858 | 19.47 | <0.001 | 0 | 1.0 | 0.341 | 0.851 |
| iqr_post_y | 1.41 | 0.245 | 33.914 | <0.001 | 10.249 | <0.001 | 0.609 | 0.656 |
| mean_trem_a | 1.037 | 0.355 | 34.677 | <0.001 | 1.947 | 0.1 | 0.934 | 0.443 |
| kur_trem_x | 1.231 | 0.293 | 3.437 | 0.064 | 1.804 | 0.125 | 0.427 | 0.789 |
| F95_post_a | 2.29 | 0.103 | 4.361 | 0.037 | 0.532 | 0.712 | 0.198 | 0.939 |
| ApEn_trem_x | 0.806 | 0.447 | 9.629 | 0.002 | 4.215 | 0.002 | 0.137 | 0.969 |
| zcr_post_y | 0.05 | 0.952 | 16.755 | <0.001 | 0 | 1.0 | -1.081 | 1 |
| median_post_a | 1.58 | 0.207 | 30.751 | <0.001 | 3.102 | 0.015 | 0.698 | 0.593 |
| iqr_trem_x | 3.391 | 0.035 | 7.537 | 0.006 | 2.05 | 0.085 | 0.343 | 0.849 |
| FD_CC_dist_x_z | 1.485 | 0.228 | 17.237 | <0.001 | 0.948 | 0.435 | 0.819 | 0.513 |
| F50_post_y | 0.611 | 0.543 | 13.841 | <0.001 | 4.862 | <0.001 | 2.136 | 0.074 |
| Power_LF_trem_x | 0.317 | 0.729 | 3.233 | 0.073 | 3.255 | 0.011 | 1.005 | 0.404 |
| range_trem_y | 0.621 | 0.538 | 16.464 | <0.001 | 12.134 | <0.001 | 0.764 | 0.548 |
| MVELO_dist_x | 5.03 | 0.007 | 11.829 | <0.001 | 12.194 | <0.001 | 0.455 | 0.769 |
| mean_post_y | 4.806 | 0.009 | 5.178 | 0.023 | 11.917 | <0.001 | 0.328 | 0.859 |
| min_post_y | 0.59 | 0.555 | 20.291 | <0.001 | 16.579 | <0.001 | 0.882 | 0.474 |
| iqr_post_x | 6.234 | 0.002 | 6.785 | 0.009 | 8.167 | <0.001 | 0.463 | 0.763 |
| kur_post_x | 1.217 | 0.297 | 6.657 | 0.01 | 2.837 | 0.023 | 0.282 | 0.89 |
| rms_trem_a | 0.253 | 0.777 | 22.949 | <0.001 | 2.598 | 0.035 | 0.798 | 0.527 |
| ApEn_post_a | 0.817 | 0.442 | 5.012 | 0.026 | 6.458 | <0.001 | 0.616 | 0.651 |
| skew_post_a | 0.981 | 0.376 | 16.017 | <0.001 | 3.468 | 0.008 | 0.685 | 0.602 |
| AREA_CC_dist_x_z | 0.229 | 0.796 | 21.152 | <0.001 | 2.189 | 0.068 | 1.096 | 0.357 |
| FD_dist_x_z | 2.931 | 0.055 | 7.317 | 0.007 | 1.018 | 0.396 | 1.194 | 0.311 |
| cov_trem_a | 2.753 | 0.065 | 14.523 | <0.001 | 1.629 | 0.164 | 0.562 | 0.69 |
| TotalPower_post_x_z | 0.188 | 0.829 | 4.258 | 0.039 | 1.226 | 0.298 | 0.496 | 0.738 |
| MFREQ_dist_x_z | 0.474 | 0.623 | 14.312 | <0.001 | 0 | 1.0 | -0.011 | 1 |
| max_post_y | 2.302 | 0.102 | 14.466 | <0.001 | 13.553 | <0.001 | 0.769 | 0.545 |
| F50_post_x | 1.259 | 0.285 | 0.975 | 0.324 | 0.631 | 0.641 | 1.372 | 0.241 |
| cov_post_a | 2.224 | 0.11 | 12.355 | <0.001 | 2.121 | 0.076 | 0.056 | 0.994 |
| range_trem_x | 0.251 | 0.778 | 1.748 | 0.187 | 2.225 | 0.064 | 0.5 | 0.735 |
| ApEn_trem_y | 0.825 | 0.439 | 10.51 | 0.001 | 3.458 | 0.008 | 0.994 | 0.409 |
| FD_CE_dist_x_z | 2.346 | 0.097 | 0.523 | 0.47 | 1.258 | 0.284 | 0.383 | 0.821 |
| mean_post_z | 1.368 | 0.256 | 1.085 | 0.298 | 1.66 | 0.156 | 1.065 | 0.372 |
| F50_post_a | 0.579 | 0.561 | 0.048 | 0.827 | 0.607 | 0.657 | 1.1 | 0.355 |
| Power_LF_trem_a | 1.381 | 0.253 | 9.566 | 0.002 | 1.45 | 0.215 | 1.072 | 0.369 |
| max_post_z | 3.408 | 0.034 | 0.884 | 0.347 | 6.177 | <0.001 | 0.531 | 0.713 |
| rms_post_a | 0.507 | 0.603 | 16.826 | <0.001 | 5.588 | <0.001 | 0.869 | 0.482 |
| F50_post_x_z | 2.256 | 0.106 | 0.006 | 0.938 | 0.157 | 0.96 | 1.421 | 0.224 |
| FREQD_post_x | 1.612 | 0.201 | 2.621 | 0.106 | 0.571 | 0.684 | 1.002 | 0.405 |
| TotalPower_post_z | 0.38 | 0.684 | 2.378 | 0.124 | 3.339 | 0.01 | 0.404 | 0.806 |
| FREQD_post_x_z | 0.951 | 0.387 | 0.258 | 0.612 | 0.194 | 0.942 | 0.084 | 0.987 |
| TotalPower_post_y | 0.043 | 0.958 | 14.926 | <0.001 | 7.7 | <0.001 | 0.091 | 0.985 |
| kur_post_a | 1.7 | 0.184 | 11.701 | <0.001 | 2.441 | 0.045 | 2.134 | 0.074 |
| max_trem_z | 1.652 | 0.193 | 1.857 | 0.174 | 2.054 | 0.084 | 0.54 | 0.706 |
| jerk_post_y | 0.375 | 0.688 | 7.077 | 0.008 | 2.766 | 0.026 | 0.632 | 0.64 |
| kur_trem_a | 0.071 | 0.932 | 0 | 1.0 | 2.942 | 0.019 | 0.856 | 0.49 |
| kur_trem_y | 0.054 | 0.947 | 0.406 | 0.524 | 2.859 | 0.022 | 1.539 | 0.188 |

PD- Parkinson’s Disease, HC- Healthy Control, PeakEnergy - Peak of energy, TotalPower- Energy between 15-3.5 Hz, rms- Root Mean Square, F50- Frequency Containing 50% of Total Power, F95- Frequency containing 95% of the total power, FRQD- Frequency of Dispersion of the Power Spectrum, MFREQ- Mean Frequency, iqr- Interquartile Range, kur- Kurtosis, zcr- Zero-Crossing Rate, ApEn- Entropy, skew- Skewness, jerk- Average jerk, MVELO- Mean velocity, FD- Fractal Dimension, FD_CE- Fractal Dimension based on the 95% Confidence Ellipse Area, min- Minimum Value, CFREQ- Centroidal Frequency, RHL- Ratio Between Power in High Frequency and Low Frequency, dist- Distance, MF- Medium Frequency (4-7Hz), VHF- Very High Frequency (>7Hz), HF- Hight Frequency (>4Hz), LF- Low Frequency (0.15-3.5Hz), trem- Tremor, post- Postural, a- Accelerometer Average Signal, x- Accelerometer Mediolateral Signal, y- Accelerometer Vertical Signal, z- Accelerometer Anteroposterior Signal, Hz- Hertz.

**Table S12.** Results from an analysis of ANOVA for repeated measurements on the features from Voice task. Features are selected based on Mann-Whitney U test that significantly differ between PD and HC at the first administration (baseline) (*P*<.05).

| Feature Name | Medication | | Diagnosis  (PD, HC) | | Repetition | | Diagnosis ×  Repetition | |
| --- | --- | --- | --- | --- | --- | --- | --- | --- |
|  | F | P | F | P | F | P | F | P |
| mean_gqc | 2.239 | 0.108 | 92.952 | <0.001 | 3.284 | 0.011 | 2.551 | 0.037 |
| p5_gqc | 0.114 | 0.892 | 49.446 | <0.001 | 1.938 | 0.101 | 0.545 | 0.703 |
| c_mean_MFCC1 | 0.492 | 0.611 | 38.813 | <0.001 | 24.89 | <0.001 | 1.836 | 0.119 |
| p95_gqc | 4.158 | 0.016 | 59.181 | <0.001 | 1.531 | 0.19 | 1.706 | 0.146 |
| std_tkeo | 0.176 | 0.838 | 2.341 | 0.126 | 3.392 | 0.009 | 1.89 | 0.109 |
| p95_tkeo | 0.543 | 0.581 | 4.843 | 0.028 | 1.932 | 0.102 | 0.874 | 0.478 |
| c_std_d11 | 1.651 | 0.193 | 3.892 | 0.049 | 0 | 1.0 | 0.786 | 0.534 |
| hnr_std | 0.836 | 0.434 | 13.227 | <0.001 | 18.399 | <0.001 | 1.165 | 0.324 |
| c_std_d12 | 1.265 | 0.283 | 4.053 | 0.044 | 0 | 1.0 | 4.556 | 0.001 |
| p75_tkeo | 3.092 | 0.046 | 1.517 | 0.218 | 2.435 | 0.045 | 2.042 | 0.086 |
| c_std_d13 | 2.121 | 0.121 | 4.497 | 0.034 | 0 | 1.0 | -2.873 | 1.0 |
| c_std_d8 | 0.72 | 0.487 | 2.962 | 0.086 | 0 | 1.0 | 1.201 | 0.308 |
| c_std_d9 | 1.669 | 0.189 | 3.243 | 0.072 | 0 | 1.0 | 0.978 | 0.418 |
| c_mean_MFCC10 | 1.361 | 0.257 | 22.916 | <0.001 | 0.796 | 0.528 | 0.586 | 0.673 |
| c_std_d10 | 2.649 | 0.072 | 0 | 1.0 | 0 | 1.0 | 2.912 | 0.02 |
| c_std_d7 | 1.583 | 0.206 | 2.291 | 0.13 | 0 | 1.0 | 2.004 | 0.091 |
| c_std_dd11 | 2.3 | 0.101 | 0 | 1.0 | 0 | 1.0 | 6.108 | <0.001 |
| c_std_d5 | 1.988 | 0.138 | 8.873 | 0.003 | 6.115 | <0.001 | 0.912 | 0.456 |
| c_std_d14 | 1.869 | 0.155 | 0 | 1.0 | 0 | 1.0 | 3.318 | 0.01 |
| c_std_d6 | 0.49 | 0.613 | 5.343 | 0.021 | 0 | 1.0 | 0.173 | 0.952 |
| c_mean_MFCC12 | 4.491 | 0.012 | 10.811 | 0.001 | 2.232 | 0.063 | 1.304 | 0.266 |
| c_std_dd5 | 2.368 | 0.095 | 0 | 1.0 | 0 | 1.0 | 6.746 | <0.001 |
| c_std_d3 | 2.208 | 0.111 | 12.446 | <0.001 | 2.773 | 0.026 | 2.114 | 0.076 |
| c_std_d4 | 1.978 | 0.139 | 6.739 | 0.01 | 3.227 | 0.012 | 0.614 | 0.653 |
| c_std_dd10 | 3.345 | 0.036 | 0 | 1.0 | 0 | 1.0 | 6.864 | <0.001 |
| c_std_dd9 | 2.126 | 0.12 | 0 | 1.0 | 0 | 1.0 | 6.709 | <0.001 |
| c_std_dd8 | 1.622 | 0.198 | 0 | 1.0 | 0 | 1.0 | 6.034 | <0.001 |
| c_std_dd12 | 1.96 | 0.142 | 0 | 1.0 | 0 | 1.0 | 6.84 | <0.001 |
| c_std_MFCC1 | 0.931 | 0.395 | 13.851 | <0.001 | 16.376 | <0.001 | 1.183 | 0.316 |
| c_std_MFCC5 | 2.659 | 0.071 | 1.814 | 0.178 | 4.874 | <0.001 | 1.613 | 0.168 |
| c_std_dd7 | 3.237 | 0.04 | 0 | 1.0 | 0 | 1.0 | 7.599 | <0.001 |
| c_std_dd13 | 3.163 | 0.043 | 0 | 1.0 | 0 | 1.0 | 6.79 | <0.001 |
| c_std_dd6 | 1.27 | 0.282 | 0 | 1.0 | 0 | 1.0 | 5.008 | <0.001 |
| DFA | 0.298 | 0.742 | 10.114 | 0.002 | 1.763 | 0.133 | 1.062 | 0.374 |
| c_mean_0th | 0.815 | 0.443 | 26.062 | <0.001 | 43.877 | <0.001 | 1.754 | 0.135 |
| c_mean_d13 | 0.43 | 0.651 | 0 | 1.0 | 0 | 1.0 | 3.693 | 0.005 |
| fm | 2.782 | 0.063 | 0.054 | 0.816 | 4.34 | 0.002 | 1.546 | 0.186 |
| c_std_dd4 | 3.189 | 0.042 | 0 | 1.0 | 0 | 1.0 | 0.546 | 0.702 |
| c_mean_MFCC7 | 1.71 | 0.182 | 13.614 | <0.001 | 1.009 | 0.401 | 1.73 | 0.14 |
| c_mean_d4 | 1.792 | 0.168 | 0 | 1.0 | 0 | 1.0 | 9.902 | <0.001 |
| shdb | 0.982 | 0.375 | 7.787 | 0.005 | 1.985 | 0.094 | 0.538 | 0.708 |
| c_std_dd3 | 4 | 0.019 | 0 | 1.0 | 0 | 1.0 | 4.832 | <0.001 |
| ApEn_f0 | 0.552 | 0.576 | 4.946 | 0.026 | 5.313 | <0.001 | 1.242 | 0.291 |
| c_std_dd14 | 2.764 | 0.064 | 0 | 1.0 | 0 | 1.0 | 10.367 | <0.001 |
| c_mean_d3 | 0.123 | 0.884 | 0 | 1.0 | 0 | 1.0 | 9.723 | <0.001 |
| c_std_MFCC11 | 0.292 | 0.747 | 0.139 | 0.709 | 3.71 | 0.005 | 0.946 | 0.436 |
| c_std_d1 | 0.414 | 0.661 | 1.638 | 0.201 | 6.838 | <0.001 | 1.692 | 0.149 |
| c_std_MFCC6 | 0.936 | 0.393 | 1.571 | 0.21 | 5.619 | <0.001 | 1.651 | 0.159 |
| c_mean_MFCC5 | 2.483 | 0.084 | 7.142 | 0.008 | 2.813 | 0.024 | 0.198 | 0.939 |
| c_mean_MFCC9 | 1.686 | 0.186 | 1.264 | 0.261 | 1.554 | 0.184 | 0.205 | 0.936 |
| c_std_MFCC9 | 0.865 | 0.422 | 0.041 | 0.84 | 4.202 | 0.002 | 0.825 | 0.509 |
| rpde | 0.274 | 0.761 | 4.674 | 0.031 | 23.492 | <0.001 | 5.175 | <0.001 |
| c_std_MFCC7 | 1.206 | 0.3 | 0.45 | 0.502 | 4.679 | <0.001 | 1.148 | 0.332 |
| c_mean_MFCC8 | 0.392 | 0.676 | 1.373 | 0.242 | 1.375 | 0.24 | 0.552 | 0.698 |
| c_std_MFCC8 | 0.084 | 0.919 | 0.154 | 0.695 | 2.876 | 0.022 | 0.919 | 0.452 |
| c_mean_MFCC6 | 0.018 | 0.982 | 0.936 | 0.334 | 2.035 | 0.087 | 1.011 | 0.4 |
| c_mean_d2 | 1.782 | 0.169 | 0 | 1.0 | 0 | 1.0 | 6.742 | <0.001 |
| c_mean_d6 | 0.515 | 0.598 | 0 | 1.0 | 0 | 1.0 | 3.16 | 0.013 |
| c_mean_MFCC3 | 1.393 | 0.249 | 1.886 | 0.17 | 7.87 | <0.001 | 6.754 | <0.001 |
| c_mean_d1 | 1.664 | 0.19 | 0 | 1.0 | 0 | 1.0 | 7.083 | <0.001 |

PD- Parkinson’s Disease, HC- Healthy Control, c_mean- Mean of the MFCCs Coefficients, log-Energy of the Signal and the First and Second Derivatives of the MFCCs, MFCC- Mel Frequency Cepstral Coefficients, c_std- Standard Deviation of the MFCCs Coefficients, gqc- Glottis Quotient Close, fm- Frequency Modulation, std - Standard Deviation, tkeo- Teager Kaiser Energy Operator, p5- 5th percentile, p75- 75th Percentile, p95- 95th Percentile, shbd- Shimmer, hnr- Harmonic to Noise Ratio, d- Delta, d-d- Delta-Delta, DFA- Detrended Fluctuation Analysis, f0- Fundamental Frequency, ApEn- Pitch Period Entropy, rpde- Recurrence Period Density Entropy, T- Period.

**Table S13.** Results from an analysis of ANOVA for repeated measurements on the features from Tapping task. Features are selected based on Mann-Whitney U test that significantly differ between PD and HC at the first administration (baseline) (*P*<.05).

| Feature Name | Medication | | Diagnosis  (PD, HC) | | Repetition | | Diagnosis ×  Repetition | |
| --- | --- | --- | --- | --- | --- | --- | --- | --- |
|  | F | *P* | F | *P* | F | *P* | F | *P* |
| numberTaps | 4.903 | 0.008 | 539.151 | <0.001 | 123.309 | <0.001 | 4.444 | 0.001 |
| max_TapInter | 0.784 | 0.457 | 296.6 | <0.001 | 4.609 | 0.001 | 1.233 | 0.294 |
| range_TapInter | 0.768 | 0.464 | 271.738 | <0.001 | 3.207 | 0.012 | 1.308 | 0.264 |
| ar2_TapInter | 0.219 | 0.803 | 159.025 | <0.001 | 6.706 | <0.001 | 2.583 | 0.035 |
| ar1_TapInter | 0.194 | 0.824 | 268.543 | <0.001 | 10.484 | <0.001 | 3.829 | 0.004 |
| sd_TapInter | 1 | 0.369 | 212.428 | <0.001 | 3.804 | 0.004 | 1.424 | 0.223 |
| buttonNoneFreq | 0.073 | 0.929 | 50.866 | <0.001 | 4.93 | <0.001 | 2.344 | 0.052 |
| mad_TapInter | 0.524 | 0.593 | 87.645 | <0.001 | 0.721 | 0.578 | 4.172 | 0.002 |
| median_DriftRight | 0.128 | 0.88 | 8.987 | 0.003 | 9.747 | <0.001 | 6.351 | <0.001 |
| mad_DriftRight | 0.167 | 0.846 | 8.9 | 0.003 | 10.605 | <0.001 | 6.353 | <0.001 |
| median_DriftLeft | 0.382 | 0.683 | 4.491 | 0.034 | 3.107 | 0.015 | 2.769 | 0.026 |
| min_TapInter | 1.49 | 0.226 | 44.616 | <0.001 | 11.437 | <0.001 | 0.388 | 0.818 |
| sd_DriftRight | 1.279 | 0.279 | 4.338 | 0.037 | 6.689 | <0.001 | 3.516 | 0.007 |
| iqr_TapInter | 1.145 | 0.319 | 53.591 | <0.001 | 1.054 | 0.378 | 2.553 | 0.037 |
| mad_DriftLeft | 0.603 | 0.548 | 2.594 | 0.108 | 11.145 | <0.001 | 6.06 | <0.001 |
| sd_DriftLeft | 0.297 | 0.743 | 4.586 | 0.032 | 3.469 | 0.008 | 2.217 | 0.065 |
| skew_TapInter | 2.064 | 0.128 | 77.596 | <0.001 | 1.117 | 0.346 | 2.171 | 0.07 |
| skew_DriftLeft | 2.606 | 0.075 | 33.216 | <0.001 | 1.021 | 0.395 | 0.684 | 0.603 |
| kur_DriftLeft | 1.888 | 0.152 | 29.675 | <0.001 | 1.196 | 0.31 | 0.414 | 0.799 |
| kur_DriftRight | 1.34 | 0.262 | 55.612 | <0.001 | 2.173 | 0.069 | 1.553 | 0.184 |
| kur_TapInter | 1.672 | 0.189 | 45.738 | <0.001 | 0.864 | 0.484 | 0.88 | 0.475 |
| cv_TapInter | 1.466 | 0.232 | 8.054 | 0.005 | 6.815 | <0.001 | 1.702 | 0.146 |
| corXY | 0.135 | 0.874 | 5.935 | 0.015 | 21.815 | <0.001 | 2.541 | 0.038 |
| tkeo_TapInter | 0.373 | 0.689 | 26.888 | <0.001 | 0.424 | 0.792 | 0.388 | 0.817 |
| cv_DriftLeft | 2.424 | 0.089 | 0.005 | 0.944 | 0.573 | 0.682 | 1.793 | 0.127 |

PD- Parkinson’s Disease, HC- Healthy Control, iqr- Interquartile Range, TapInter- Tap Interval, buttonNoneFreq: Frequency of Tapping Outside the Button, numberTaps- Number of Taps, DriftRight- Right Drift, corXY- Correlation of X and Y Positions, DriftLeft- Left Drift, mad- Median Absolute Deviation, min- Minimum, max- Maximum, skew- Skewness, kur- Kurtosis, teko- Teager-Kaiser Energy Operator, cv- Coefficient, Sd- Standard Deviation, ar (1-2)- Coefficient of an Autoregressive Model at Lag (1-2).

### Reference

1.	Mirelman A, Heman T, Yasinovsky K, Thaler A, Gurevich T, Marder K, et al. Fall risk and gait in Parkinson’s disease: the role of the LRRK2 G2019S mutation. Mov Disord 2013 Oct 7;28(12):1683–1690. PMID: 24123150

2.	Sejdić E, Lowry KA, Bellanca J, Redfern MS, Brach JS. A comprehensive assessment of gait accelerometry signals in time, frequency and time-frequency domains. IEEE Trans Neural Syst Rehabil Eng 2014 May;22(3):603–612. PMID: 23751971

3.	Zhan A, Little MA, Harris DA, Abiola SO, Dorsey ER, Saria S, et al. High Frequency Remote Monitoring of Parkinson’s Disease via Smartphone: Platform Overview and Medication Response Detection. arXiv 2016 Jan 5;

4.	Arora S, Venkataraman V, Zhan A, Donohue S, Biglan KM, Dorsey ER, et al. Detecting and monitoring the symptoms of Parkinson’s disease using smartphones: A pilot study. Parkinsonism Relat Disord 2015 Jun;21(6):650–653. PMID: 25819808

5.	Weiss A, Sharifi S, Plotnik M, van Vugt JPP, Giladi N, Hausdorff JM. Toward automated, at-home assessment of mobility among patients with Parkinson disease, using a body-worn accelerometer. Neurorehabil Neural Repair 2011 Dec;25(9):810–818. PMID: 21989633

6.	San-Segundo R, Torres-Sánchez R, Hodgins J, De la Torre F. Increasing robustness in the detection of freezing of gait in parkinson’s disease. Electronics 2019 Jan 22;8(2):119.

7.	Bächlin M, Plotnik M, Roggen D, Maidan I, Hausdorff JM, Giladi N, et al. Wearable assistant for Parkinson’s disease patients with the freezing of gait symptom. IEEE Trans Inf Technol Biomed 2010 Mar;14(2):436–446. PMID: 19906597

8.	Palmerini L, Rocchi L, Mellone S, Valzania F, Chiari L. Feature selection for accelerometer-based posture analysis in Parkinson’s disease. IEEE Trans Inf Technol Biomed 2011 May;15(3):481–490. PMID: 21349795

9.	Martinez-Mendez R, Sekine M, Tamura T. Postural sway parameters using a triaxial accelerometer: comparing elderly and young healthy adults. Comput Methods Biomech Biomed Engin 2012;15(9):899–910. PMID: 21547782

10.	Prieto TE, Myklebust JB, Hoffmann RG, Lovett EG, Myklebust BM. Measures of postural steadiness: differences between healthy young and elderly adults. IEEE Trans Biomed Eng 1996 Sep;43(9):956–966. PMID: 9214811

11.	Tsanas A, Little MA, McSharry PE, Ramig LO. Nonlinear speech analysis algorithms mapped to a standard metric achieve clinically useful quantification of average Parkinson’s disease symptom severity. J R Soc Interface 2011 Jun 6;8(59):842–855. PMID: 21084338

12.	Prince J, Andreotti F, De Vos M. Multi-Source Ensemble Learning for the Remote Prediction of Parkinson’s Disease in the Presence of Source-Wise Missing Data. IEEE Trans Biomed Eng 2019;66(5):1402–1411. PMID: 30403615

13.	GitHub - Sage-Bionetworks/mPower-sdata: scripts used for preparation of Nature Scientific Data submission and release of mPower data [Internet]. [cited 2020 Aug 5]. Available from: https://github.com/Sage-Bionetworks/mPower-sdata/

14.	Synapse | Sage Bionetworks [Internet]. [cited 2020 Aug 5]. Available from: https://www.synapse.org/#!Synapse:syn4993293/files/
